# Metagenomic analysis reveals extreme complexity of *Plasmodium* spp. infections in high transmission in West Africa

**DOI:** 10.1101/2025.04.29.25326533

**Authors:** Mun Hua Tan, Oscar Bangre, Cecilia A. Rios-Teran, Kathryn E. Tiedje, Samantha L. Deed, Qi Zhan, Fathia Rasyidi, Mercedes Pascual, Patrick O. Ansah, Karen P. Day

## Abstract

Mixed-species, mixed-strain *Plasmodia* infections are known to occur in humans in malaria endemic areas. To date, the true extent of this complexity has not been explored in high-burden countries of sub-Saharan Africa. Here we take a metagenomic lens to infections obtained by sampling variable blood volumes from residents living in high, seasonal transmission in northern Ghana. We identified significantly higher prevalence of *Plasmodium* spp. and inter-/intra-species complexity in larger blood volumes. Overall, malaria infections displayed high levels of metagenomic complexity comprising single-, double-, and triple-species infections with varying levels of complexity for *P. falciparum, P. malariae, P. ovale curtisi, and P. ovale wallikeri.* We present evidence of individuals with greater susceptibility to highly-complex infections that cannot be explained by age or location. The implications of these findings to malaria epidemiology and control are illustrated by a geographic scaling exercise to district and region levels in Ghana.

## 1. Introduction

Hematozoan parasites represent a taxonomic group of eukaryotic microorganisms that infect a broad range of hosts. The most studied of this group are the causative agents of malaria, *Plasmodium* spp., a taxon that has been shown to be highly diverse^1^. The definitive hosts of malaria parasites, including humans, lizards, birds, and non-human primates, are frequently infected with multiple diverse species and strains, e.g.^2–8^. Despite this diversity, malaria is often too simplistically considered a disease caused by a single infectious agent, due in the case of humans to the dominating prevalence of a single species of *Plasmodium falciparum* that causes the major burden of infection globally. Often overlooked is the extreme genomic diversity of *P. falciparum* strains and other *Plasmodium* spp. in a host and how they may interact^5,9^.

The most recent World Health Organisation (WHO) Malaria Report shows that 95% of malaria cases occur in sub-Saharan Africa, with most found in eleven high-burden countries in Africa, including Ghana^10^. In these high-transmission endemic areas, individuals develop non-sterilising immunity protecting against clinical disease but not infection due to the extreme genomic diversity of *Plasmodium* spp., especially of *P. falciparum* with less known about the other species infecting humans^11–17^. These infections are typically asymptomatic and key to sustaining transmission^18^. Surveillance of the asymptomatic reservoir in humans has typically observed a large gradient in parasite density for *P. falciparum*, with most of these being submicroscopic or low-density infections with densities significantly lower than observed in clinical infections (Fig. S1). Whilst it is known that individuals harbour infections with mixed species and strains^19,20^, the extent of this within-host complexity in the asymptomatic reservoir has never been explored.

Here we present a metagenomic approach to study malaria in high transmission, given that malaria infection in a person comprises a community of parasites. Current reliance on whole genome sequencing with limitations in methods to phase genomes in highly-complex infections has forced the field to view infections simplistically by examining dominant parasites whilst ignoring rare species or strains with potential for emergence. In contrast, a key aspect of metagenomic experimental design is ensuring deep sampling to capture most of the complexity of a community, as employed in environments as diverse as human microbiome and space microbiology^21–23^. Similarly, this must be considered when designing malaria surveillance and genetic studies in high transmission, where the end game is to eliminate the species or detect the emergence of drug-resistant parasites. It is currently unknown if the routine sampling of a small blood volume from dried blood spot (DBS) cuttings in epidemiological surveys is reflective of actual metagenomic complexity per person in high transmission.

We set out to explore the impact of deep blood volume sampling on the metagenomic complexity of *Plasmodium* spp. in 188 afebrile human hosts in four age groups, living in high transmission in Bongo District, Ghana. We report significantly higher prevalence of *Plasmodium* spp. with deeper sampling compared to DBS and also observed higher *P. falciparum* complexity defined as multiplicity of infection (MOI). This contributed to increased richness in the population whilst low genetic similarity among isolates was maintained. We showed that complexity of malaria infections in the Sahel region of Ghana comprises single-, double-, and triple-species infections with varying MOI levels per species and identified individuals more susceptible to highly-complex infections. Our findings determined that both broad and deep sampling of a reservoir is crucial for accurate representation of the parasite population in an endemic area, requiring us to view complexity cumulatively in a metagenomic sense, not only of the dominant *P. falciparum* species but also of the minor *Plasmodium* spp. species inclusively.

## 2. Results

To measure infection complexity across species, we define here terms used in this study (Fig. 1). Sex is an obligatory part of the *Plasmodium* spp. life-cycle and constant outcrossing occurs in nature, especially in high transmission^24,25^. Therefore, unlike in bacteriology where strains are defined as clonal entities, this study defines “strains” as genetically-diverse parasites that are distinct either by antigen (for *P. falciparum* and *P. ovale* spp.) or by neutral markers (microsatellites, for *P. malariae*). The number of different strains per person is represented by “multiplicity of infection” (MOI). “Metagenomic complexity” further summarises MOI across the combination of species as a measure of overall within-host complexity.

**Fig. 1.**
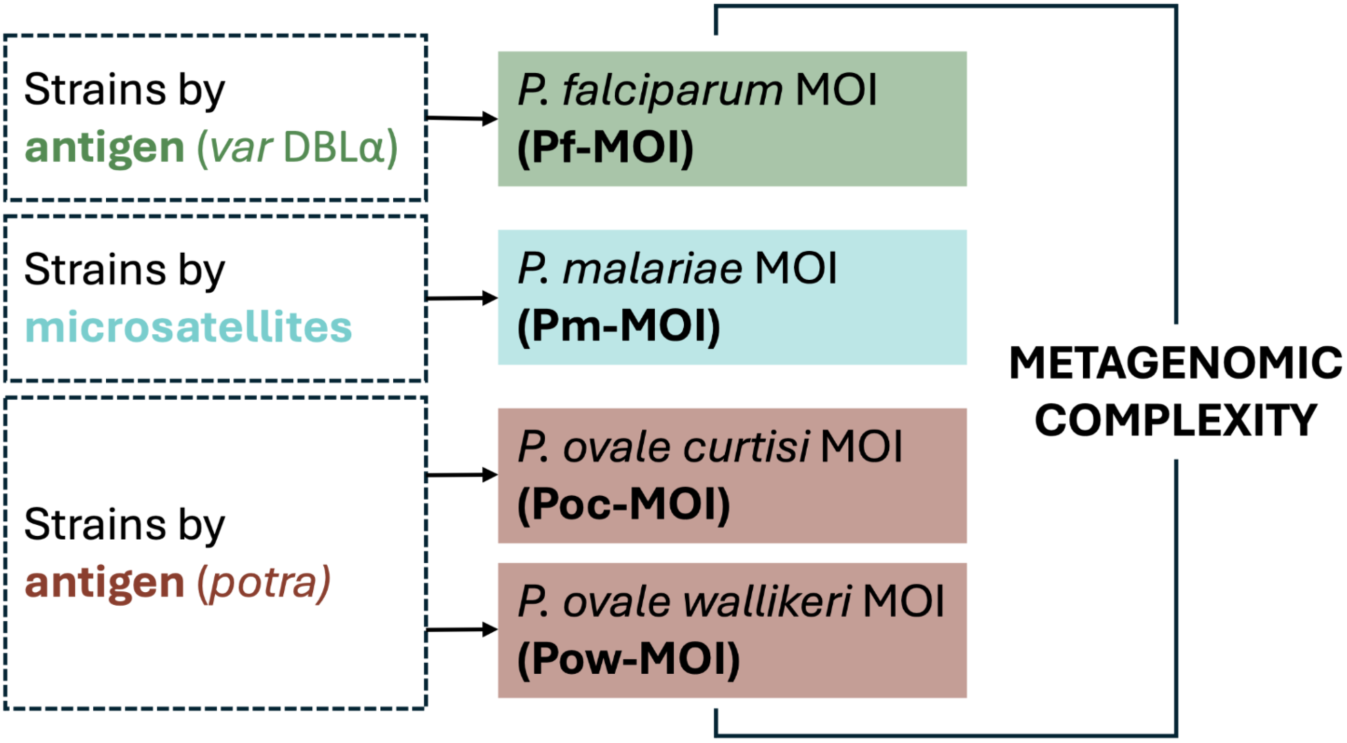
Defining terminologies in this study. We define “strains” of malaria parasites as genetically-diverse parasites that are distinct either antigenically (for *P. falciparum* and *P. ovale* spp.) or by genotypes based on neutral microsatellites (for *P. malariae*). “Multiplicity of infection” (MOI) represents the number of different strains per person. “Metagenomic complexity” summarises MOI across the combination of species as a measure of overall within-host complexity.

### 2.1 Demography of participants and parasitological characteristics of infections

This study was conducted in the Bongo District (Vea/Gowrie catchment area) in the Upper East Region of northern Ghana, characterised by seasonal malaria transmission^26,27^. Based on the WHO “A Framework for Malaria Elimination”^28^, this area is categorised as “high transmission” where *P. falciparum* microscopic prevalence was ≥35% at baseline in 2012^26,27^. A study population of 188 individuals (i.e. “isolates”) with age ranges 6-90 years were sampled in November 2020 (Tables S1, S2). Children <5 years were receiving seasonal malaria chemoprevention (SMC) and excluded. Of the 188 isolates, 75.5% were adolescents (11-20 years) and adults (≥21 years), generally excluded in current malaria surveillance where the target is measuring disease burden^29^. Only 17.6% of the 188 isolates were found to be microscopically-positive for *Plasmodium* spp., predominantly of *P. falciparum*, including mixed infections. We also identified microscopy-negative isolates with positive *P. falciparum* histidine-rich protein 2 (PfHRP2) measurements.

### 2.2 Higher prevalence of *Plasmodium* spp. multi-species infections in larger pRBC volumes

Overall, *Plasmodium* spp. prevalence was underestimated when DBS cuttings or small blood volumes were sampled. Based on *18S rRNA*, parasite detection levels were significantly higher in 100μL packed red blood cells (pRBC) relative to those in DBS, by factors of 1.32× for *P. falciparum*, 1.93× for *P. malariae*, and 2.50× for *P. ovale* spp. (Fig. 2). An independent *var*coding protocol corroborated *P. falciparum* findings, indicating 1.36× detection in 100μL compared to 1μL-pRBC (Table S3). *P. vivax* remained absent from infections regardless of blood volume sampling. Increased prevalence of double- and triple-species infections in 100μL-pRBC relative to DBS was also observed (Fig. 2). Parasite prevalence relies on counting the proportion of human hosts infected but does not capture within-host parasite population sizes; we explore this within-host complexity and diversity in the subsequent sections.

**Fig. 2.**
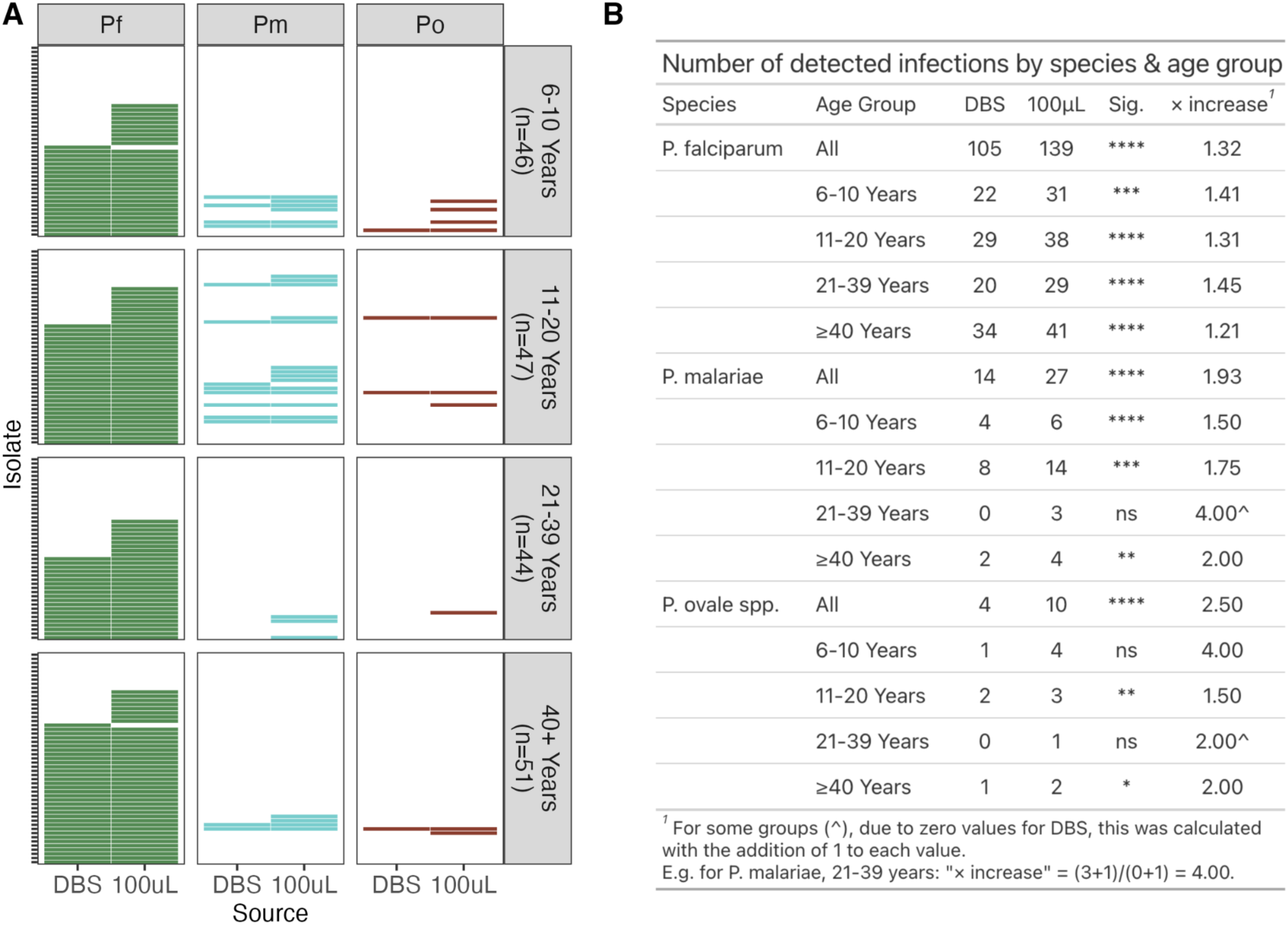
Relative to sampling of DBS, sampling of larger pRBC volumes yielded increased prevalence of *Plasmodium* spp. infections. Data shown for *Plasmodium* spp. detection by species-specific *18S rRNA* PCR on DBS and 100μL-pRBC for *N=*188 isolates, stratified by age (6-10 years (*n*=46), 11-20 years (*n*=47), 21-39 years (*n*=44), and ≥40 years (*n*=51)). All infections were negative for *P. vivax* and therefore excluded from this figure. (A) Presence or absence of *P. falciparum*, *P. malariae*, and *P. ovale* spp. as indicated by “Pf”, “Pm”, and “Po” respectively. (B) Number of detected infections by species and age group. Fisher’s exact test was used to assess significance in differences between DBS and 100uL-pRBC, with symbols *, **, ***, and **** representing p-value significance levels of 0.05, 0.01, 0.001, and 0.0001, respectively.

### 2.3 Intra-species complexity of *P. falciparum*

#### 2.3.1 Greater intra-species complexity of *P. falciparum* in larger pRBC volumes

Many isolates had greater *P. falciparum* strain complexity with deeper blood sampling (Fig. 3A, Table S4). Using a fingerprinting method (*var*coding) shown to more accurately estimate *P. falciparum* MOI (Pf-MOI) in high-transmission areas^30,31^, 101 isolates had Pf-MOI ≥ 1 across all four pRBC volumes. Distributions of Pf-MOI were significantly different (*p-value < 0.0001*), with the largest increase observed between 1μL- and 100μL-pRBC with median Pf-MOI of 1 and 3, respectively (*p-value < 0.0001*) (Fig. 3B). Concordance was high for Pf-MOI from 50μL and 100μL, suggesting near-saturation with these larger volumes (Fig. S2) though Pf-MOI distributions remained significantly different (*p-value < 0.01*) (Fig. 3B). Most DBLα types found in 1μL-pRBC were also present in the larger volumes (Fig. S3, S4). High concordance in Pf-MOI was estimated between 40 repeat isolates at all volumes and Pf-MOI distributions between repeats were not significantly different, supporting a robust approach in this study (Fig. S5).

**Fig. 3.**
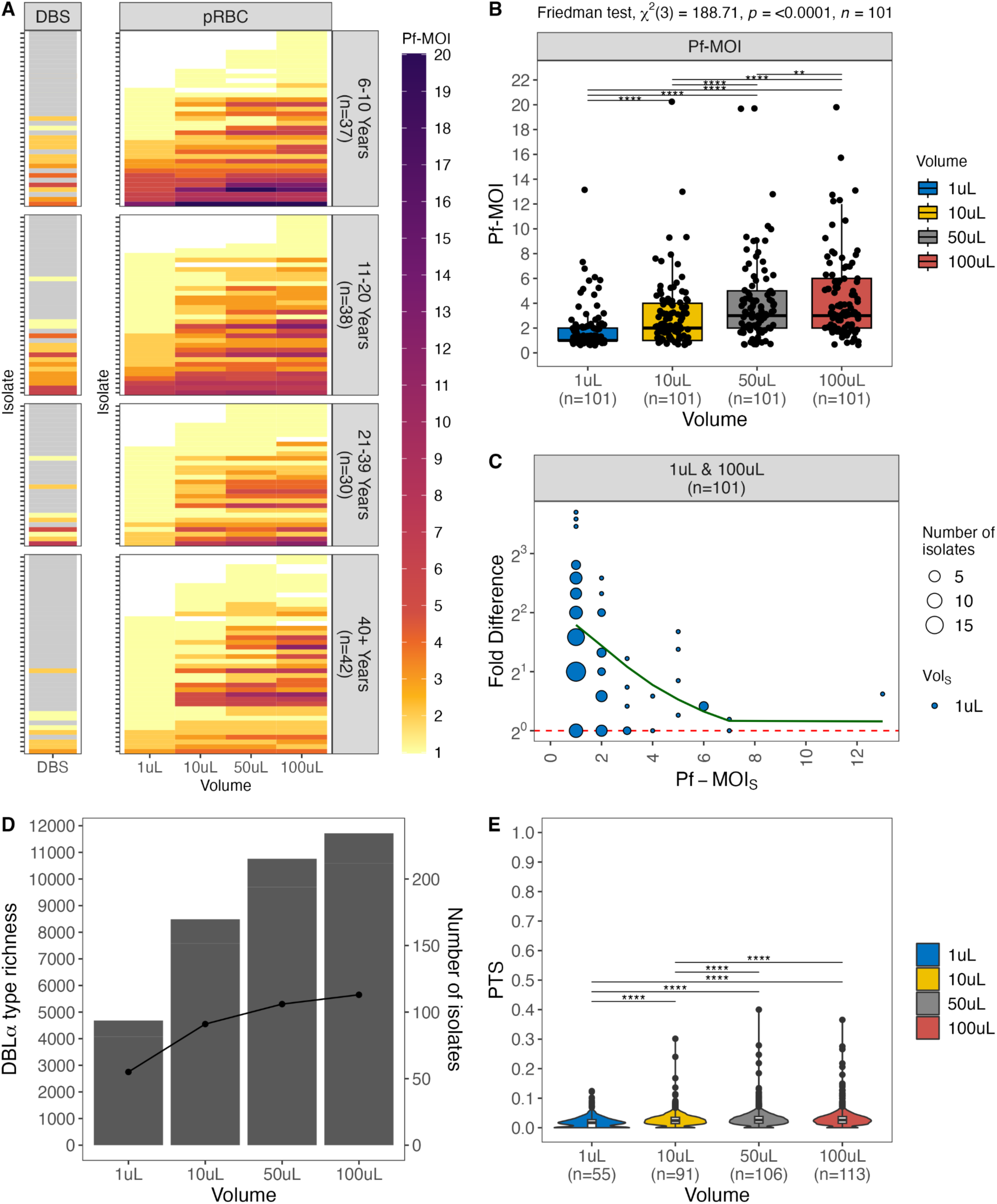
Higher levels of *P. falciparum* Pf-MOI were detected in larger pRBC volumes. (A) Shown here are only 147 isolates with Pf-MOI ≥ 1 in at least one pRBC volume, stratified by age (6-10 years (*n*=37), 11-20 years (*n*=38), 21-39 years (*n*=30), and ≥40 years (*n*=42)). For DBS, isolates in grey were not *var*coded as these were negative by microscopy. (B) Boxplots are shown for isolates with Pf-MOI ≥ 1 for all four pRBC volumes (*n=*101). Significance of pairwise pRBC comparisons was inferred with pairwise Wilcoxon signed rank tests, with levels of significance adjusted using the “holm” correction. (C) Non-linear relationship between Pf-MOI fold differences (FD*Pf-MOI* in log scale) and the Pf-MOI measured from 1μL-pRBC volumes (Pf-MOI*S*), shown for isolates with Pf-MOI ≥ 1 for all four pRBC volumes (*n=*101). Red dashed line indicates no difference in Pf-MOI (FD*Pf-MOI* = 1) while green solid line shows the smooth curve fitted with a generalised additive model. (D) Population-level metrics were estimated for isolates with Pf-MOI ≥ 1 with isolate repertoire size ≥ 20. Differences in DBLα type richness and number of infected isolates are shown, represented by bars and points, respectively. (E) Differences in genetic similarity between isolate repertoires by pairwise-type sharing (PTS), where most distributions were significantly different, with the exception of 50μL *vs* 100μL. Boxes within each violin show the median and interquartile ranges, values available in Table S6. Statistical analysis was conducted with the Kruskal-Wallis analysis of variance, followed by Dunn’s test for multiple comparisons. In all plots, significance symbols *, **, ***, and **** represent p-value significance levels of 0.05, 0.01, 0.001, and 0.0001, respectively.

#### 2.3.2 Non-linear relationships between Pf-MOI fold differences and initial Pf-MOI

For isolates with Pf-MOI = 0 in a pRBC volume but Pf-MOI ≥ 1 in a different volume, these Pf- MOI values mostly differed by one (Fig. S6). For the 101 isolates with Pf-MOI ≥ 1 across all four pRBC volumes, relationships between fold differences in Pf-MOI (FD*Pf-MOI*) and the Pf- MOI in the smaller compared pRBC volumes (Pf-MOI*S*) were non-linear (Fig. 3C, Fig. S7). Relatively small FD*Pf-MOI* values were observed for MOI comparisons between 50μL- *vs* 100μL- pRBC, suggesting again that sampling was almost saturated at these volumes sufficient to capture the complexity of *P. falciparum* multiclonal infections in our study population. These non-linear correlations can be considered for use to computationally adjust Pf-MOI values estimated for DBS samples in large-scale field surveillance. A power analysis based on proportions of Pf-MOI > 1 isolates in each pair of 1μL *vs* larger pRBC volume determined that there was sufficient data for comparison of different pRBC volumes when all isolates were considered, but larger sample sizes are needed for age-stratified analyses (Table S5).

#### 2.3.3 Increased *P. falciparum* complexity added to richness while infections remained largely unrelated

Our observations can be used to evaluate potential differences in epidemiological and population genetics metrics when variable blood volumes are sampled for a same set of isolates. Increased DBLα type richness was observed with larger pRBC sampling volumes (Fig. 3D) while DBLα type sharing between isolate repertoires remained low for all four pRBC volumes, indicating minimal genetic overlap (Fig. 3E, Table S6). Thus, not only did the deeper sampling of volume detect greater complexity of strains within infections in the form of higher Pf-MOI, this additional complexity contributed newly-detected diversity in the population such that we did not find clonal or highly-related parasites between isolates, as evidenced from rising cumulative diversity (Fig. S8).

#### 2.3.4 Shallow broad sampling *vs* deep restricted sampling: is one approach enough for malaria surveillance?

We also had DBS samples from 1,809 individuals from our larger cross-sectional survey in Bongo collected during the same surveyed time point. Of these, 263 were positive for asymptomatic *P. falciparum* infections by microscopy. When comparing DBLα types recovered from DBS of 263 asymptomatic isolates and 100μL-pRBC from 113 isolates, only 32.1% of the total 21,222 DBLα types was seen in combined DBS and 100μL-pRBC samples (Fig. S9, Table S7). Deep vertical sampling of the population using 100μL-pRBC volumes recovered 23.1% of the total DBLα types that were not recovered in broad sampling of DBS of the same Bongo population at relatively shallow depths. The remainder of 44.8% of total DBLα types was recovered by DBS sampling but not seen in the pRBC sampling. Finding subsets of DBLα types exclusively from different sample sources suggests that both sampling approaches are necessary and complementary to more accurately reflect true population- level metrices.

### 2.4 Inter- and intra-species complexity of *Plasmodium* spp

We report on infection complexity for three minor species *P. malariae* (Pm-MOI), *P. ovale curtisi* (Poc-MOI), and *P. ovale wallikeri* (Pow-MOI) in 100μL-pRBC samples of isolates previously identified as positive by *18S rRNA*. For 26 *P. malariae* isolates, microsatellite data revealed a median Pm-MOI of 2 [range: 1-4]. Of the ten *P. ovale* spp. isolates, there were eight *P. ovale curtisi*, one *P. ovale wallikeri*, and one double-species infection based on the *P. ovale* spp. tryptophan-rich antigen gene (*potra*), with fragment sizes reporting Poc-MOI and/or Pow-MOI of 1.

For 146 isolates with any MOI data, the summation of estimated MOI across all four species is expressed as metagenomic complexity (Fig. 4A), showing single-, double-, or triple-species infections (Fig. 4B). Whilst the one triple Pm-Poc-Pow infection reflected an absence of *P. falciparum* (i.e. *var*coding data, Pf-MOI = 0), *P. falciparum* was detected in this isolate with *18S rRNA* PCR, suggesting a possible quadruple-species infection. The absence of *P. falciparum* in the two Pm-only infections was supported by *18S rRNA* PCR (i.e., not detected) and *var*coding data (i.e., Pf-MOI = 0).

**Fig. 4.**
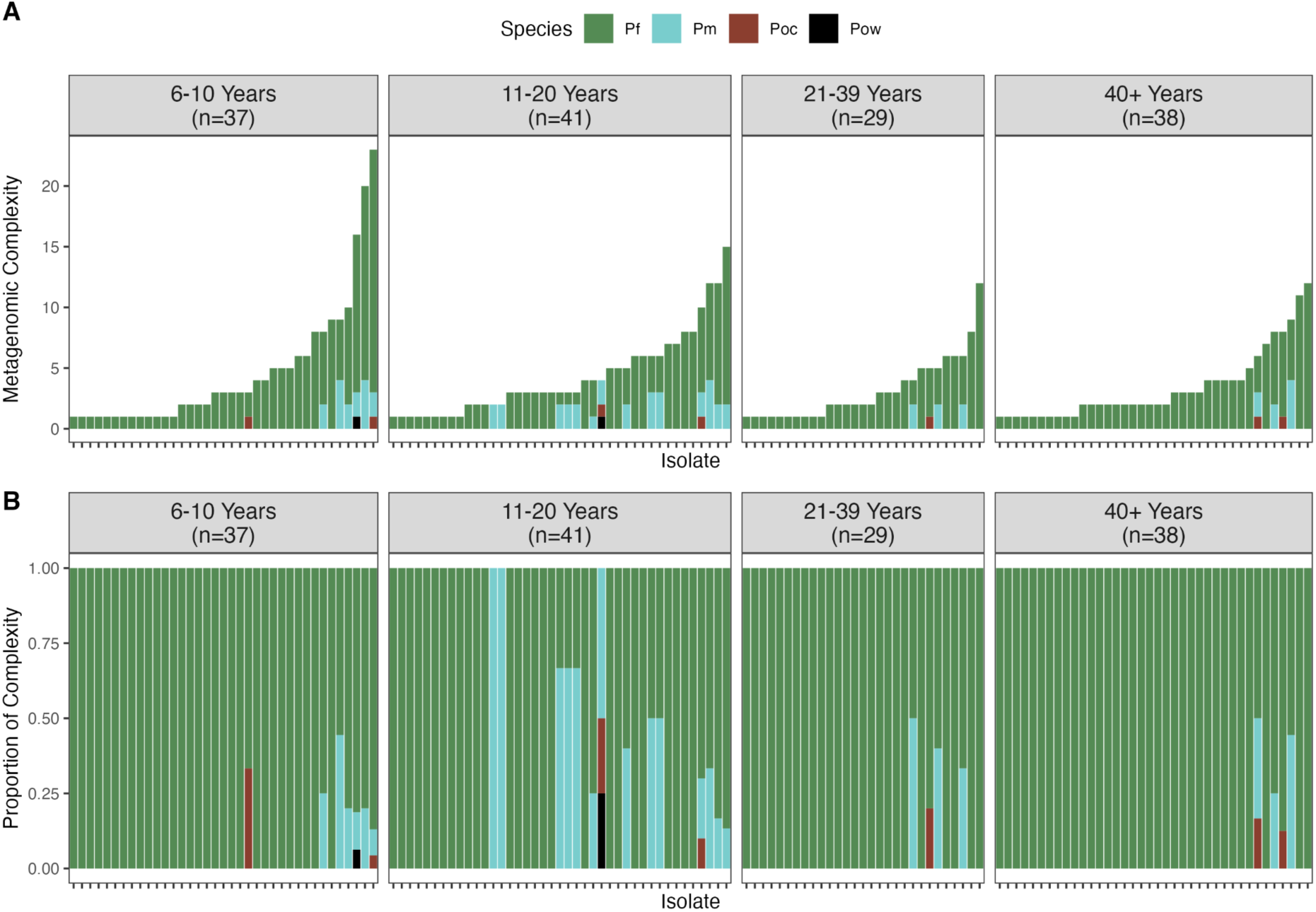
Malaria infection complexity comprising of single-, double-, and triple-species infections with varying MOI levels. Isolates are ordered in increasing metagenomic complexity, i.e. total MOI from four combined *Plasmodium* spp. species. (A) Metagenomic complexity calculated from the summation of estimated MOI across all four species of *P. falciparum* (Pf), *P. malariae* (Pm), *P. ovale curtisi* (Poc), and *P. ovale wallikeri* (Pow). (B) Proportion of complexity representing the proportion of MOI contributed per species.

Extreme metagenomic complexity with summed MOI from 10-23 was seen in a subset of individuals in every age group (Fig. 4A), occurring at a prevalence of 5.85% in this survey of 188 individuals. Most of these were in the groups of children and adolescents. These individuals had as many as 7-20 different *P. falciparum* antigenic strains, 2-4 *P. malariae* genotypes by microsatellites, and/or 1 each of *P. ovale curtisi*/*P. ovale walikeri* antigenic strains by *potra*. It is unclear why extreme complexity was found in these individuals, comprising nine males and two females. Inspection of association between metagenomic complexity and human host or spatial characteristics did not yield any notable findings (Table S8).

### 2.5 Impact on malaria surveillance and elimination

By comparing DBS or 1uL-pRBC to deeper sampling of 100uL-pRBC, this study quantified the underestimation of prevalence and metagenomic complexity in current protocols employing small blood volumes. Exemplified below is the application of these quantified factors to adjust estimates from the larger survey of DBS samples of 1,809 individuals in Bongo, of which 1,455 were aged ≥6 years.

The Malaria Atlas Project (MAP) reports standardised rapid diagnostic tests and microscopy-based *P. falciparum* prevalence of 8.9% in Bongo, based on the proportion of children aged 2-10 years in a defined year^32^. Employing molecular methods on DBS samples, we estimated higher *P. falciparum* prevalence in the same age group of children aged 2-10 years (34.6%) and even higher prevalence when individuals of all ages were inspected (49.5%) (Fig. 5A). Following adjustments to account for deeper volume sampling, a 65.6% *P. falciparum* prevalence was estimated for Bongo. This study additionally made available prevalence data for minor *Plasmodium* species currently unavailable in MAP.

**Fig. 5.**
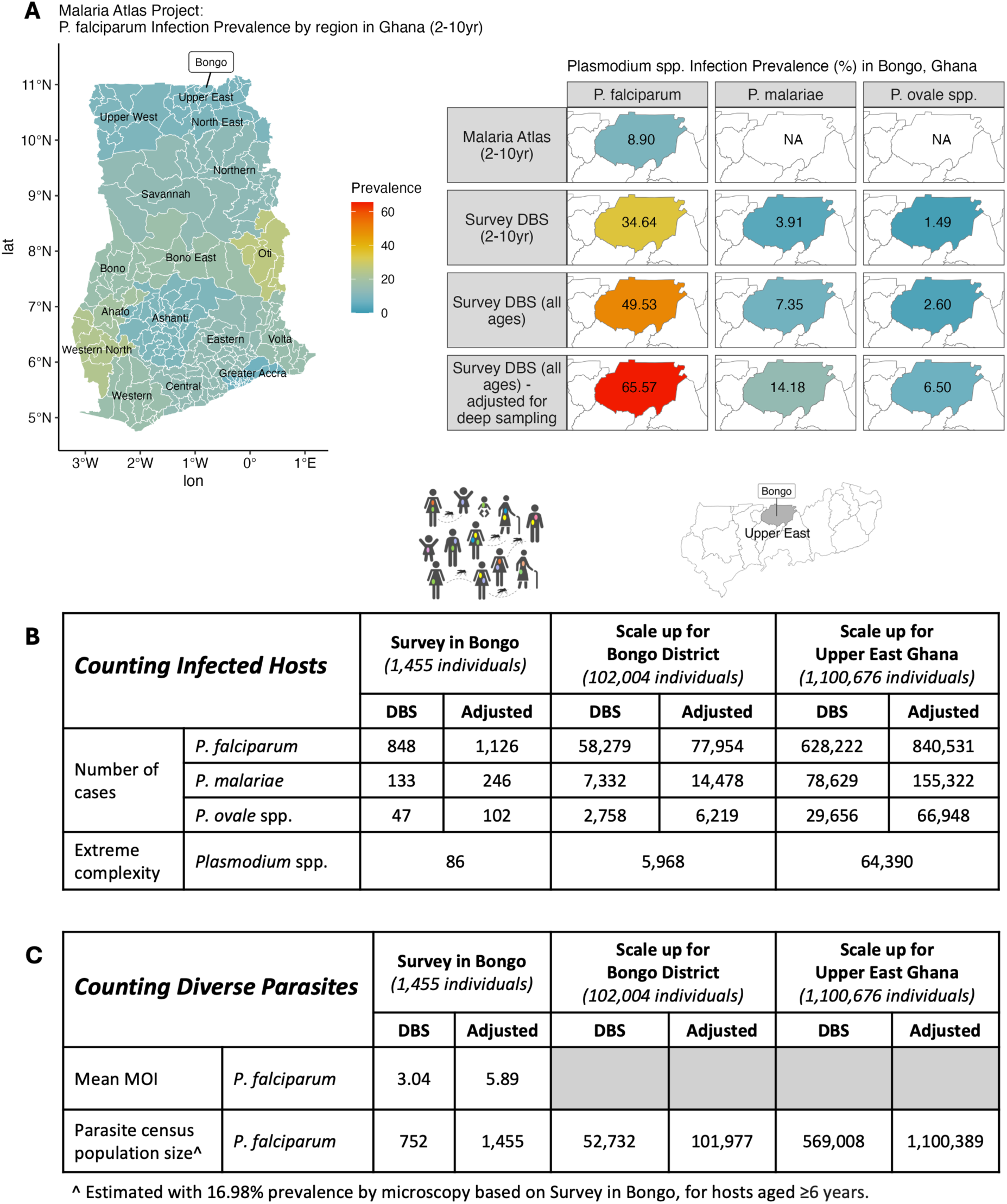
Impact of underestimated complexity on the large reservoir in the Bongo District and the Upper East Region in Ghana. In this figure, “Adjusted” represents adjusted estimates to account for deeper volume sampling (i.e. 100μL). (A) LEFT: Malaria Atlas Project (MAP) report of *P. falciparum* infection prevalence by region in Ghana in children 2-10 years. RIGHT: Comparison of *P. falciparum* prevalences in Bongo estimated by MAP in children 2-10 years, by *18S rRNA* in our larger DBS survey in children 2-10 years, in our larger DBS survey in all ages, and adjusted for deeper volume sampling in all ages. Also shown are prevalences for minor species *P. malariae* and *P. ovale* spp. that are not available in MAP. (B) Impact on metrics that count infected hosts. Prevalence and adjusted prevalence estimated from the larger DBS survey in individuals ≥6 years were scaled up to approximate the number of malaria cases in the Bongo DBS survey, Bongo District, and the Upper East Region in Ghana, detailed in Table S9. Similarly, the numbers of heavily-infected individuals with extreme metagenomic complexity in these regions were approximated with the 5.85% prevalence observed in this study. (C) Impact on metrics that count diverse parasites. Similarly, mean Pf-MOI and adjusted mean Pf-MOI estimated from the larger DBS survey in individuals ≥6 years were scaled up to approximate the parasite census population size in the Bongo DBS survey, Bongo District, and the Upper East Region in Ghana without age stratification.

To illustrate the relevance of these findings to malaria surveillance and elimination, we scaled up prevalence and *P. falciparum* complexity data to the district and regional levels in Bongo and Upper East Ghana, respectively, based on a 2021 Population and Housing census by the Ghana Statistical Service (GSS)^33,34^. Scale up of difference in prevalence of *P. falciparum*, *P. malariae*, and *P. ovale* spp. (Fig. 5B) predicted that tens of thousands of *Plasmodium* spp. infections would be missed in Bongo District from using DBS, and hundreds of thousands would be missed in the wider Upper East Region in Ghana (Table S9). In a population, thousands of these individuals are expected to be heavily infected with highly-complex infections, representing a substantial reservoir. This study provides a sampling frame to look for these individuals for further exploration.

Whilst parasite prevalence and clinical incidence count the number of infected human hosts (Fig. 5A and B), other metrics such as MOI and parasite census population size count the number of diverse parasite genomes in infections (Fig. 5C). A mean Pf-MOI of 3.04 was calculated from *var*coding of DBS samples from 247 individuals microscopy-positive for *P. falciparum*. Subsequently, mean Pf-MOI was adjusted to 5.89 to account for deeper volume sampling, resulting in larger parasite census population sizes in Bongo District and the Upper East Region in Ghana.

## 3. Discussion

Our metagenomic approach of sampling variable blood volumes for *Plasmodium* spp. species in Bongo, Ghana highlighted the complexity of infections in this area of West Africa. This endemic area is typical of high seasonal transmission in the Sahel so our results can be broadly translated to this high-burden region of West Africa, even those experiencing intense malaria control^35,36^. This contemporary deep dive into *Plasmodium* spp. infection complexity, post- multiple interventions^37^ found *P. falciparum* as the dominant species with minor species *P. malaria*, *P.ovale curtisi*, and *P. ovale wallikeri* present, generally at submicroscopic levels. *P. vivax* was undetected in the Bongo population, likely due to the significant protection of Duffy-negativity against sustained endemic *P. vivax* infections in West African human hosts^38^. Scaling up to district and regional level, we predicted that most individuals of all ages are infected with *Plasmodium* spp. and current DBS surveys are significantly underestimating the prevalence of minor species infections.

Our report of single-, double-, and triple-species infections comprising varying MOI levels per species shows that highly complex infections are common in this asymptomatic reservoir in Ghana. Of note, we observed a subset of individuals with extreme metagenomic complexity of infection in all age classes. Whether susceptibility to extreme mixed infections is due to behavioural, host genetic, immunological, or environmental factors warrants further investigation. This observation is reminiscent of “wormy” people observed in helminth ecology/epidemiology studies^39^ and predicted by anopheline preferential biting behaviour^39^. These heavily-infected individuals warrant further investigation.

Given the elevated diversity and frequent multiclonal infections in the malaria reservoir in high-transmission areas, metrics of parasite prevalence and clinical incidence that use the infected human host as the unit of measurement do not capture the complexity of infections in the asymptomatic reservoir. We have illustrated the impact of scaling prevalence of infected hosts based on deeper sampling in Bongo District and the larger Upper East Region of Ghana. Scaling up on within-host diversity must consider the observed non-linear relationship between fold differences in Pf-MOI and the Pf-MOI in the smaller compared pRBC volumes, with the obvious implication that DBS estimates cannot be corrected by a single factor of difference. Instead, such a factor must be calculated using a measured distribution, where adjustments are mostly needed for isolates with Pf-MOI in the 1 to 3 range. Larger sample sizes stratified by age, gender, and relevant heterogeneities to an endemic area will be necessary to explore these non-linear relationships further as will Bayesian approaches^32^. This type of scale-up of a study to multiple spatial sites counting both infected hosts and genetically-diverse parasites per host has broad applications for geographic information system (GIS) mapping, such as for the Malaria Atlas Project^32^. Our findings make the case that both broad and deep sampling of all ages in sentinel sites is needed to achieve accurate representation of an asymptomatic reservoir.

Malaria elimination in Africa requires a focus on the reservoir, characterised by low-density subclinical infections. Yet, there have been limited investigations of these low-density subclinical infections of *Plasmodium* species in general^40–43^, with the majority of current molecular methods developed for high-density, clinical samples. In this context, our study serves as a warning of the extent of the malaria problem in terms of prevalence and diversity in the reservoir of infection in all ages in high-burden, high-transmission areas in Africa. Data presented also point to the need for a change in perspective and approaches in malaria genomic surveillance, so that we view malaria infections holistically as the complex infections that they are rather than the examination of each species individually^17,44–47^. In doing so, we can better study the impact of transmission-reducing interventions such as indoor residual spraying (IRS) and vaccines on the reservoir^37^.

Additionally, metagenomics offers a way to study emerging drug resistance in rare strains and minor species conferred through current treatment regimens optimised for *P. falciparum*^48–50^. With prolonged SMC and widespread use of antimalarials to *P. falciparum* in communities, the probability of drug resistance is likely to increase silently in *P. malariae* and *P. ovale* spp., pointing to the need for deep sampling of the reservoir. Such mutations have already been found with investigators using whole genome amplification and large volumes of blood from clinical cases where parasitaemias are significantly higher^51^. Metagenomics further allows us a means to examine complex interactions among species (e.g.^5^) revealed by epidemiological patterns as well as emergence of minor species with relapse capability. To achieve this, accurate measurement of infection complexity is crucial by taking on board metagenomic approaches to sampling. Broadly, our results are also relevant to those doing surveillance of Haemosporidian parasites of other hosts infected with multiple species and genotypes.

## 4. Methods

Detailed methods are available in Supplementary Data 1.

### 4.1 Study area and population

This study was conducted in the Bongo District (Vea/Gowrie catchment area) in the Upper East Region of northern Ghana. Informed consent was obtained from key stakeholders and the local community in Bongo District. Members of the local community were trained as field workers directly involved in liaising with the community and in the collection of data. At the time of this survey, the population had undergone malaria control interventions, including long-lasting insecticidal nets (LLINs), three rounds of indoor residual spraying (IRS) (2013- 2015), and five consecutive years (2016-2020) of seasonal malaria chemoprevention (SMC) administered at monthly intervals during the malaria season to children between the ages of 3-59 months (i.e. <5 years old)^30^. This study was reviewed and approved by the ethics committees at the Navrongo Health Research Centre, Ghana (NHRC IRB-131) and The University of Melbourne, Australia (HREC 21649).

### 4.2 Estimation of WB and pRBC volume equivalent in DBS

The typical protocol in the field involves collecting 3-4 dried blood spots (DBS) per filter paper^52^. From these small DBS, two sections are cut (∼5mm × 5mm each) for gDNA extraction. We approximated that two 5mm × 5mm DBS cuttings contains the equivalent of ∼6-7μL of whole blood (Supplementary Data 2). Given the average expected proportion of RBCs at ∼40%, our typical DBS cuttings would thus have the equivalent of ∼2.4-2.8μL packed red blood cells (pRBC), with some variation due to a person’s hermatocrit levels.

### 4.3 Parasitological measurements

Parasitological measurement methods were performed as previously published^37,53^. Briefly, parasite densities were counted against 200 white blood cells (WBC) on 10% Giemsa-stained thick film blood smears and examined under oil immersion of 100-fold magnification. Final parasite densities (parasites/mL of blood) were calculated by averaging two independent readings, assuming an average WBC count of 8,000/μL of blood. Parasite species were identified using a 100-fold magnification of thin film smears and categorised based on morphology.

### 4.4. Sampling of venous whole blood and dried blood spot samples

#### 4.4.1. Whole blood (WB)

200 individuals with ages ranging 6-90 years were sampled. Children <5 years receiving SMC were excluded. Approximately 5mL of venous whole blood samples (WB) were collected per person and transported to the Navrongo Health Research Centre, where they were processed within 2-4 hours after collection. These WB samples were centrifuged to separate the WB into components of pRBC, white blood cells, and plasma. All three components were harvested individually and stored at -80°C.

#### 4.4.2. Dried blood spots (DBS)

At the same time as the venous whole blood collection, DBS were also collected onto filter paper (3MM Whatman) for the same participants and for a larger subset of the population (see ^30^ for detailed methods on DBS collection).

### 4.5. Extraction of genomic DNA from pRBC and DBS

Of the 200 individuals (i.e. “isolates”), a total of 12 were excluded (Table S1). Nine were excluded as these could not be matched accurately to epidemiological data. The remaining three isolates were excluded as these were later confirmed to be symptomatic (i.e. febrile and microscopically-positive for *P. falciparum*). Genomic DNA was extracted from four volumes of pRBC (1μL, 10μL, 50μL, 100μL) from the remaining 188 afebrile isolates. Genomic DNA was eluted in 50μL buffer AE. In this study, a “sample” refers to a DBS or pRBC volume per isolate, hence there can be multiple samples per isolate.

Two 5mm × 5 mm sections were cut from DBS and gDNA was extracted as previously published^26^. Genomic DNA was eluted in 50μL buffer AE.

### 4.6. Measurement of plasma PfHRP2 concentrations

Plasma PfHRP2 concentration was determined using the Quantimal CELISA kit (TM, Cellabs). In this enzyme-linked immunosorbent assay, wells are coated with a primary monoclonal antibody to PfHRP2, sample is applied and wells are washed, then probed with a secondary anti-Pf antibody conjugated to horseradish peroxidase (HRP) for detection at 450nm.

### 4.7. Detection of *Plasmodium* spp. using *18S rRNA* species-specific PCR

*Plasmodium* species detection was performed on the DBS and 100μL-pRBC samples (*N=*188 isolates). To detect the presence of different *Plasmodium* species, previously published protocols with modifications were used^26,54^. A nested polymerase chain reaction targeting the 18S ribosomal RNA gene (*18S rRNA*) was performed^26^ to identify *Plasmodium* species in DBS and pRBC samples^25^. The different *Plasmodium* spp. were determined to be present based on band size (*P. falciparum*: 205bp, *P. malariae*: 144bp, *P. ovale* spp.: 800bp, *P. vivax*: 120bp).

### 4.8. Genotyping of *P. malariae* using microsatellites

Of the 188 isolates, 26 were identified as *P. malariae*-infected isolates through species-specific PCR. The 100µL-pRBC sample for each of these isolates was genotyped at 12 neutral microsatellite markers using previously published primers^55,56^. A semi-nested PCR protocol was adapted for low-density infections (Rios-Teran et al., ‘In preparation’). Four pools were prepared per isolate for fragment analysis by capillary electrophoresis. Allele sizes were measured by comparing with the size standard LIZ500. Only alleles greater than one third of the dominant allele were scored to avoid stutter peaks. Automated binning was performed using *tandem*^57^.

### 4.9. Identification and genotyping of *P. ovale* spp. using *potra*

Of the 188 isolates, 10 were identified as *P. ovale* spp.-infected isolates through species-specific PCR. The 100µL-pRBC sample for each of these isolates was genotyped for *P. ovale* spp. identification (i.e., *P. ovale curtisi* or *P. ovale wallikeri*) and subsequently genetic diversity. This semi-nested PCR protocol involves *P. ovale* spp. specific amplification of a size-polymorphic fragment of the tryptophan-rich antigen gene (*potra*)^16,58^ with modifications for low-density infections (Rios-Teran et al., ‘In preparation’). Visualised on 2% agarose gel, the species *P. ovale curtisi* or *P. ovale wallikeri* was present if a band was seen around 400-600bp.

### 4.10. Targeted amplicon sequencing of *P. falciparum* DBLα tags with *var*coding

The *var*coding method has been shown to be appropriate for MOI estimation in high transmission, outperforming SNP-based barcodes^31,59^. This method utilises targeted amplicon sequencing of *var* DBLα type sequences encoding a Duffy-binding-like domain of *Plasmodium falciparum* erythrocyte membrane protein 1 (PfEMP1). For DBS samples, *var*coding was performed on microscopy-positive isolates using primers and protocols previously described^30^.

For pRBC samples, a modified protocol was applied, detailed in Supplementary Data 1. Four pRBC volumes for 188 isolates were *var*coded, including those that appear negative by microscopy or species-specific PCR. Forty isolates were randomly selected as repeats to assess the reproducibility. Laboratory strains were included as positive controls. Samples of varying pRBC volumes of a same isolate and repeats were sequenced on the Illumina MiSeq platform (2 × 300bp) in the same sequencing pool and run.

### 4.11. Processing DBLα tags into DBLα types

An established bioinformatics workflow^60,61^ consisting of a suite of pipelines was used to generate DBLα tags from raw Illumina paired-end reads (https://github.com/UniMelb-Day-Lab/tutorialDBLalpha). DBLα tag sequence data included DBLα tags from pRBC generated in this study, combined with DBLα tags from an interrupted time-series study in Bongo, involving one pilot and eight cross-sectional surveys conducted between 2012 to 2020 (i.e. Malaria Reservoir Study). These surveys involved mostly asymptomatic isolates^26,30,62^ with small proportions of symptomatic and clinical isolates (unpublished). DBLα data cleaning^60^, clustering^60^ at a 96% nucleotide identity threshold^63^, and classification into ups groups^64^ was performed with published pipelines (Supplementary Data 1).

### 4.12. Estimation of isolate repertoire size and multiplicity of infection (Pf-MOI)

The isolate repertoire size represents the number of unique DBLα types in an isolate. Pf-MOI represents the estimated number of unique parasite genomes in an isolate. For samples with ≥20 DBLα types, Pf-MOI in each sample was estimated using a Bayesian approach (prior=“uniform”, aggregate=“pool”)^30^. Samples with ≥1 DBLα types but <20 DBLα types were assigned MOI=1.

### 4.13. Estimation of fold difference (FD*Pf-MOI*) and model fitting

Differences in Pf-MOI were expressed as fold differences (FD):

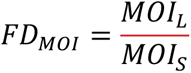

where MOI*S* and MOI*L* are the Pf-MOI of the smaller (S) and larger (L) volume samplings, respectively, of a same isolate. A generalised additive model (GAM) was fit to the data (FD*Pf- MOI* and Pf-MOI*S*) with penalised smoothing parameters selected by REML (*gam* function in mgcv v1.9-1^65^).

### 4.14. Estimation of genetic similarity or overlap between isolate repertoires

To ensure robust analyses exploring genetic similarity, we evaluated only samples with ≥ 20 DBLα types (i.e. isolate repertoire size ≥ 20). Genetic similarity of repertoires of two isolate repertoires is generally represented by the pairwise type sharing metric (PTS)^63^. Specifically:

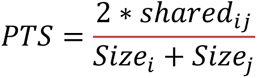

where shared*ij* is the shared number of DBLα types between isolate repertoires of samples *i* and *j*, and Size*i* and Size*j* are the isolate repertoire sizes of samples *i* and *j*, respectively. A value of 0 indicates the absence of sharing between two samples while a value of 1 indicates completely identical DBLα repertoires.

Genetic similarity between isolate repertoires from two different pRBC volumes was assessed in a directional manner relative to the isolate repertoire size of a reference sampled volume (i.e. denominator)^60^:

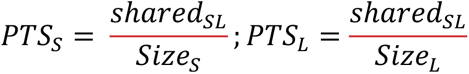

where shared*SL* is the shared number of DBLα types between isolate repertoires of smaller (S) and larger (L) volume samplings of a same isolate, and Size*S* and Size*L* are the isolate repertoire sizes of smaller and larger volume samplings, respectively, of a same isolate (i.e. Vol*S* and Vol*L*). High PTS*S* and low PTS*L* values indicate that DBLα types identified in the smaller volume are also present in the larger volume and that there are additional DBLα types identified in the larger volume but not found in the smaller volume.

### 4.15. Scaling up prevalence, Pf-MOI, and parasite census population size to larger regions in Bongo and Upper East Ghana

Prevalence, mean MOI, and parasite census population size^18^ values were estimated from DBS samples of a larger surveyed 1,455 individuals aged ≥6 years. These values were further adjusted to obtain estimates if isolates were sampled more deeply at 100μL (Supplementary Data 1). Further scaling up to the larger population sizes of Bongo and Upper East Region of Ghana provided an approximation of the number of infections or parasites underestimated in these regions. Given the exclusion of children ≤ 5 years from this study, population size data was split for 0-9 years with a 6:4 ratio into two age groups 0-5 years and 6-9 years. Relevant to our study. The final age-structured population size data as follows:

- Bongo District: 102,004 [≥6 years]; 12,166 [6-9 years]; 28,494 [10-19 years]; 35,135 [20-39 years]; 26,209 [≥40 years].
- Upper East Region: 1,100,676 [≥6 years]; 133,700 [6-9 years]; 304,105 [10-19 years]; 378,958 [20-39 years]; 283,913 [≥40 years].

Map of regions and districts in Ghana in Fig. 5A was drawn with Global ADMinistrative area (GADM) data downloaded for Ghana (v4.1)^66^.

### 4.16. Statistical analysis

Differences in the number of *Plasmodium* spp. species detected between DBS and 100µL- pRBC were analysed with Fisher’s exact test. The Friedman test compared MOI distributions for paired data of four pRBC volumes. Pairwise Wilcoxon signed rank tests with “holm” correction compared MOI data of paired pRBC volumes or repeats. Lin’s concordance correlation coefficient (CCC) estimated the agreement of MOI values between pRBC volumes for a same isolate or between two repeats of a same isolate. The Kruskal-Wallis Rank Sum test compared PTS distributions between repeats for each pRBC volumes, with Dunn’s test for multiple comparisons. Association of variables with metagenomic complexity was tested using the Mann-Whitney U test (sex, village), the Kruskal-Wallis test (occupation, section), and Spearman’s correlation (age, haemoglobin, temperature). Tests with *p-value* or adjusted *p-value* ≤ 0.05 were considered significant. Through power analysis, we determined the minimum sample sizes to achieve enough statistical power of 0.80 (α=0.05, alternative=“two.sided”) for future correction by host age groups.

## Data availability

Primer sequences, DBLα type sequences, and data tables underlying results are available on GitHub (https://github.com/mh-tan/Metagenomic_Complexity_Plasmodium).

## Acknowledgements

We thank the participants, communities, and the Ghana Health Service in Bongo District, Ghana for their participation in this study. We further thank the field teams in Bongo for their technical assistance in the field, as well as the laboratory personnel at the Navrongo Health Research Centre for their expertise and for undertaking the sample collections and parasitological assessments. Negative control of Melbourne plasma was kindly provided by Prof Stephen Rogerson’s lab. This research was supported by The University of Melbourne’s Research Computing Services and the Petascale Campus Initiative. This research was supported by the University of Melbourne Early Career Researcher Grant (TA503450) awarded to M-H.T, and the National Institute of Allergy and Infectious Diseases, National Institutes of Health through the joint NIH-NSF-NIFA Ecology and Evolution of Infectious Disease award R01-AI149779 to K.P.D and M.P. Salary support for M-H.T., O.B., S.L.D., and K.E.T. was provided by R01-AI149779.

## Author Contributions

M-H.T. and K.P.D. conceptualised the study and designed the experiments. O.B. and P.O.A. led field collection of microscopy and blood samples in Ghana. M-H.T., C.A.R-T., and S.L.D. performed gDNA extractions. S.L.D. measured PfHRP2 from plasma. C.A.R-T. and F.R. performed species-specific PCR. M-H.T. and K.E.T. performed *var*coding of pRBC and DBS, respectively. C.A.R-T. performed genotyping of *P. malariae* and *P. ovale* spp.. M-H.T. analysed the data. M-H.T. and Q.Z. performed fitting of data. M.P. supervised Q.Z.. M-H.T. and K.P.D. interpreted the results and wrote the manuscript. M-H.T., K.P.D., and M.P. funded the study. All authors reviewed the manuscript.

