## Supplementary Figures and Tables for "Metagenomic analysis reveals extreme complexity of *Plasmodium* spp. infections in high transmission in West Africa"

### Supplementary Information

Additional figures and tables

**Fig. S1. Microscopic *P. falciparum* densities were significantly lower in asymptomatic isolates compared to clinical isolates during the same epidemiological survey.** Parasite densities in clinical isolates are ~55x that of asymptomatic isolates, with median densities of 29,600 parasites/µL (*N*=93, IQR: 15,720-44,000) and 520 parasites/µL (*N*=295, IQR: 240-2,120) for clinical and asymptomatic, respectively. Distributions are significantly different by Wilcoxon rank sum test (*p-value < 0.001*).

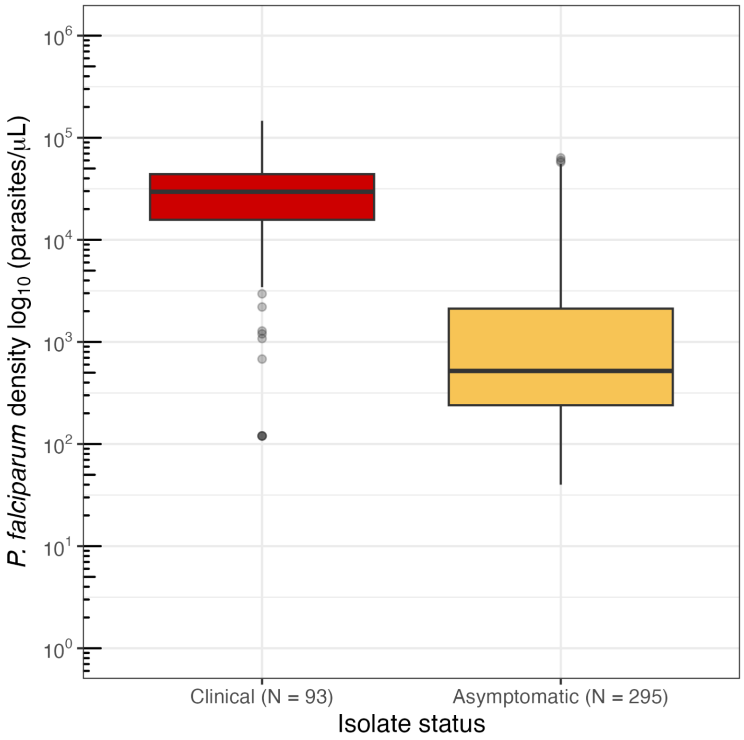

#### Table S1. Dataset sizes following exclusion of isolates in the analysis workflow.

| Parameters | Remaining number of isolates |
|---|---|
| **I.** Isolates sampled for pRBC | 200 |
| **II.** Isolates with no pRBC duplicates | 193 (96.5%)* |
| **III.** Isolates with no DBS duplicates | 192 (96.0%)* |
| **IV.** Isolates with matching epidemiological data | 191 (95.5%)* |
| **V.** Isolates found to be not symptomatic | 188 (94.0%)* |
| **VI.** Isolates with repertoire size ≥ 1 DBLα types in any of the four pRBC volume (*var*coding)^ | 147 (78.2%) |
| 1μL+ | 105 (71.4%) |
| 10μL+ | 119 (81.0%) |
| 50μL+ | 131 (89.1%) |
| 100μL+ | 142 (96.6%) |
| **VII.** Isolates with repertoire size ≥ 20 DBLα types in any of the four pRBC volume (*var*coding)^ | 113 (60.1%) |
| 1μL+ | 55 (48.7%) |
| 10μL+ | 91 (80.5%) |
| 50μL+ | 106 (93.8%) |
| 100μL+ | 113 (100.0%) |

* Data reflect No. (% [n/N]) of 200 isolates sampled in (I).
^ Data reflect No. (% [n/N]) of remaining isolates in (V).
+ Data reflect No. (% [n/N]) of remaining isolates in (VI or VII).

#### Table S2. Demography of participants and parasitological characteristics of *Plasmodium* spp. infections.

| Characteristic | | This study |
|---|---|---|
| **Sample size*** | | |
| **All** | | 188 |
| **Age groups** | 6-10 years | 46 (24.5%) |
| | 11-20 years | 47 (25.0%) |
| | 21-39 years | 44 (23.4%) |
| | ≥40 years | 51 (27.1%) |
| **Sex** | Female | 100 (53.2%) |
| | Male | 88 (46.8%) |
| **Village** | Vea | 89 (47.3%) |
| | Gowrie | 99 (52.7%) |
| **Microscopy-positive^** | | |
| **All** | | 33 (17.6%) |
| **Age groups** | 6-10 years | 9 (27.3%) |
| | 11-20 years | 12 (36.4%) |
| | 21-39 years | 4 (12.1%) |
| | ≥40 years | 8 (24.2%) |
| **Sex** | Female | 22 (66.7%) |
| | Male | 11 (33.3%) |
| **Village** | Vea | 16 (48.5%) |
| | Gowrie | 17 (51.5%) |
| ***Plasmodium* spp. median density†** | | |
| **All** | | 720 [320-2880] |
| **Age groups** | 6-10 years | 840 [360-3720] |
| | 11-20 years | 1,100 [490-5760] |
| | 21-39 years | 240 [130-330] |
| | ≥40 years | 520 [120-1600] |
| **Sex** | Female | 520 [320-3900] |
| | Male | 960 [280-1800] |
| **Village** | Vea | 520 [240-1880] |
| | Gowrie | 900 [320-3850] |

**Abbreviations:** IQR, interquartile range
* Data reflect No. (% [n/N]) of participants sampled.
^ Data reflect No. (% [n/N]) of participants sampled that were positive for *Plasmodium* spp., predominantly infected with *P. falciparum* (including mixed *P. falciparum* infections).
† Median parasite density for the microscopically-positive *Plasmodium* spp. isolates, predominantly *P. falciparum* isolates (including mixed *P. falciparum* infections) (parasites/μl [IQR]).

**Table S3. Proportions of *P. falciparum* detection by *var*coding (N=188 isolates).** Detection is shown for the four pRBC volumes at a minimum isolate repertoire size thresholds of 1 and 20 DBLα types. E.g. "N-Y-Y-Y" indicates positive detection in 10, 50, and 100µL volumes only.

| Detection code (1-10-50-100µL) | Minimum isolate repertoire size | Number of Isolates | Proportion of Isolates |
|---|---|---|---|
| N-N-N-N | 1 | 41 | 21.8 |
| N-N-N-Y | 1 | 12 | 6.4 |
| N-N-Y-N | 1 | 2 | 1.1 |
| N-N-Y-Y | 1 | 11 | 5.9 |
| N-Y-N-Y | 1 | 2 | 1.1 |
| N-Y-Y-N | 1 | 2 | 1.1 |
| N-Y-Y-Y | 1 | 13 | 6.9 |
| Y-N-N-Y | 1 | 2 | 1.1 |
| Y-N-Y-Y | 1 | 1 | 0.5 |
| Y-Y-Y-N | 1 | 1 | 0.5 |
| Y-Y-Y-Y | 1 | 101 | 53.7 |

**Table S4. Microscopic detection and *P. falciparum* multiplicity of infection (Pf-MOI) categorised by host age group.** Data reflect No. (% [n/N]) of isolates in each category. Pf-MOI ≥ 1 defined for isolates with repertoire size ≥ 1.

| Host age group (years) | MOI | TOTAL | | | | Microscopy negative (-) | | | | Microscopy positive (+) | | | |
|---|---|---|---|---|---|---|---|---|---|---|---|---|---|
| | | **1µL** | **10µL** | **50µL** | **100µL** | **1µL** | **10µL** | **50µL** | **100µL** | **1µL** | **10µL** | **50µL** | **100µL** |
| All | 0 | 83 (44.1) | 69 (36.7) | 57 (30.3) | 46 (24.5) | 82 (52.9) | 69 (44.5) | 57 (36.8) | 46 (29.7) | 1 (3.0) | 0 (0.0) | 0 (0.0) | 0 (0.0) |
| | ≥ 1 | 105 (55.9) | 119 (63.3) | 131 (69.7) | 142 (75.5) | 73 (47.1) | 86 (55.5) | 98 (63.2) | 109 (70.3) | 32 (97.0) | 33 (100.0) | 33 (100.0) | 33 (100.0) |
| | **TOTAL** | **188** | | | | **155** | | | | **33** | | | |
| 6-10 | 0 | 21 (45.7) | 18 (39.1) | 16 (34.8) | 9 (19.6) | 20 (54.1) | 18 (48.6) | 16 (43.2) | 9 (24.3) | 1 (11.1) | *NA* | *NA* | *NA* |
| | ≥ 1 | 25 (54.3) | 28 (60.9) | 30 (65.2) | 37 (80.4) | 17 (45.9) | 19 (51.4) | 21 (56.8) | 28 (75.7) | 8 (88.9) | 9 (100.0) | 9 (100.0) | 9 (100.0) |
| | **TOTAL** | **46** | | | | **37** | | | | **9** | | | |
| 11-20 | 0 | 17 (36.2) | 17 (36.2) | 16 (34.0) | 9 (19.1) | 17 (48.6) | 17 (48.6) | 16 (45.7) | 9 (25.7) | 0 (0.0) | 0 (0.0) | 0 (0.0) | 0 (0.0) |
| | ≥ 1 | 30 (63.8) | 30 (63.8) | 31 (66.0) | 38 (80.9) | 18 (51.4) | 18 (51.4) | 19 (54.3) | 26 (74.3) | 12 (100.0) | 12 (100.0) | 12 (100.0) | 12 (100.0) |
| | **TOTAL** | **47** | | | | **35** | | | | **12** | | | |
| 21-39 | 0 | 23 (52.3) | 18 (40.9) | 14 (31.8) | 15 (34.1) | 23 (57.5) | 18 (45.0) | 14 (35.0) | 15 (37.5) | 0 (0.0) | 0 (0.0) | 0 (0.0) | 0 (0.0) |
| | ≥ 1 | 21 (47.7) | 26 (59.1) | 30 (68.2) | 29 (65.9) | 17 (42.5) | 22 (55.0) | 26 (65.0) | 25 (62.5) | 4 (100.0) | 4 (100.0) | 4 (100.0) | 4 (100.0) |
| | **TOTAL** | **44** | | | | **40** | | | | **4** | | | |
| ≥40 | 0 | 22 (43.1) | 16 (31.4) | 11 (21.6) | 13 (25.5) | 22 (51.2) | 16 (37.2) | 11 (25.6) | 13 (30.2) | 0 (0.0) | 0 (0.0) | 0 (0.0) | 0 (0.0) |
| | ≥ 1 | 29 (56.9) | 35 (68.6) | 40 (78.4) | 38 (74.5) | 21 (48.8) | 27 (62.8) | 32 (74.4) | 30 (69.8) | 8 (100.0) | 8 (100.0) | 8 (100.0) | 8 (100.0) |
| | **TOTAL** | **51** | | | | **43** | | | | **8** | | | |

**Fig. S2. Agreement of Pf-MOI values for pairs of pRBC volumes sampled, indicated by Lin's concordance correlation coefficient (CCC) values.** Plots are shown for isolates with Pf-MOI ≥ 1 (and with isolate repertoire size ≥ 1) for all four pRBC volume. Pf-MOI$_S$ and Pf-MOI$_L$ represent *P. falciparum* MOI of the smaller and larger pRBC volumes, respectively, in a pairwise comparison. The strongest concordance in Pf-MOI was observed for 50µL and 100µL pRBC volumes whereas the lowest concordance in Pf-MOI was observed for 1µL and 100µL pRBC volumes.

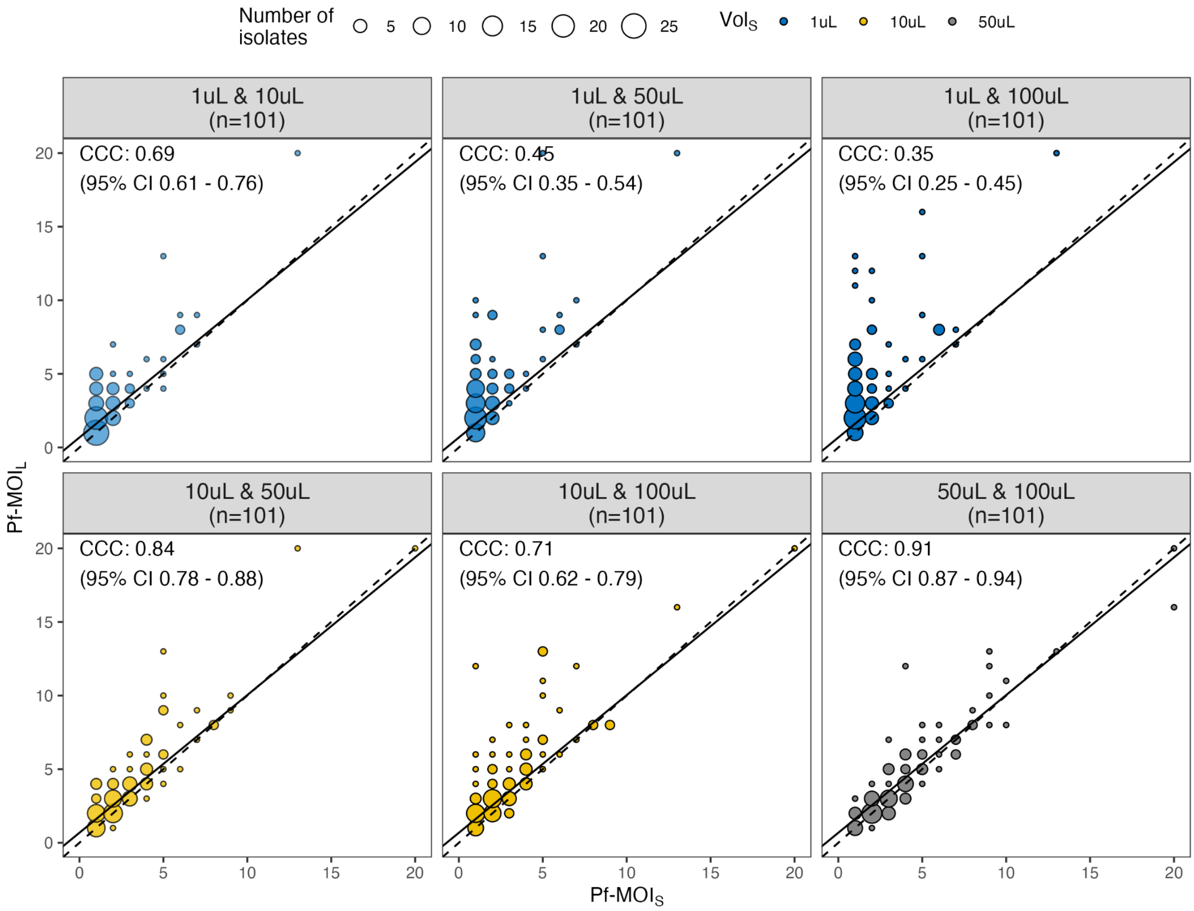

**Fig. S3. Directional genetic similarity between pairwise sampling of pRBC volumes, coloured by permutation of *P. falciparum*.** Plots are shown for isolates with Pf-MOI ≥ 1 for all four pRBC volumes (and with isolate repertoire size ≥ 20). $PTS_S$ and $PTS_L$ represent directional genetic similarity levels calculated relative to repertoire sizes of the smaller and larger pRBC volumes, respectively, in a pairwise comparison. **(A)** Each data point represents an isolate, coloured by its permutation of *P. falciparum* detection in the four pRBC volumes. **(B)** Distributions of directional PTS values are coloured by the smaller volume ($Vol_S$).

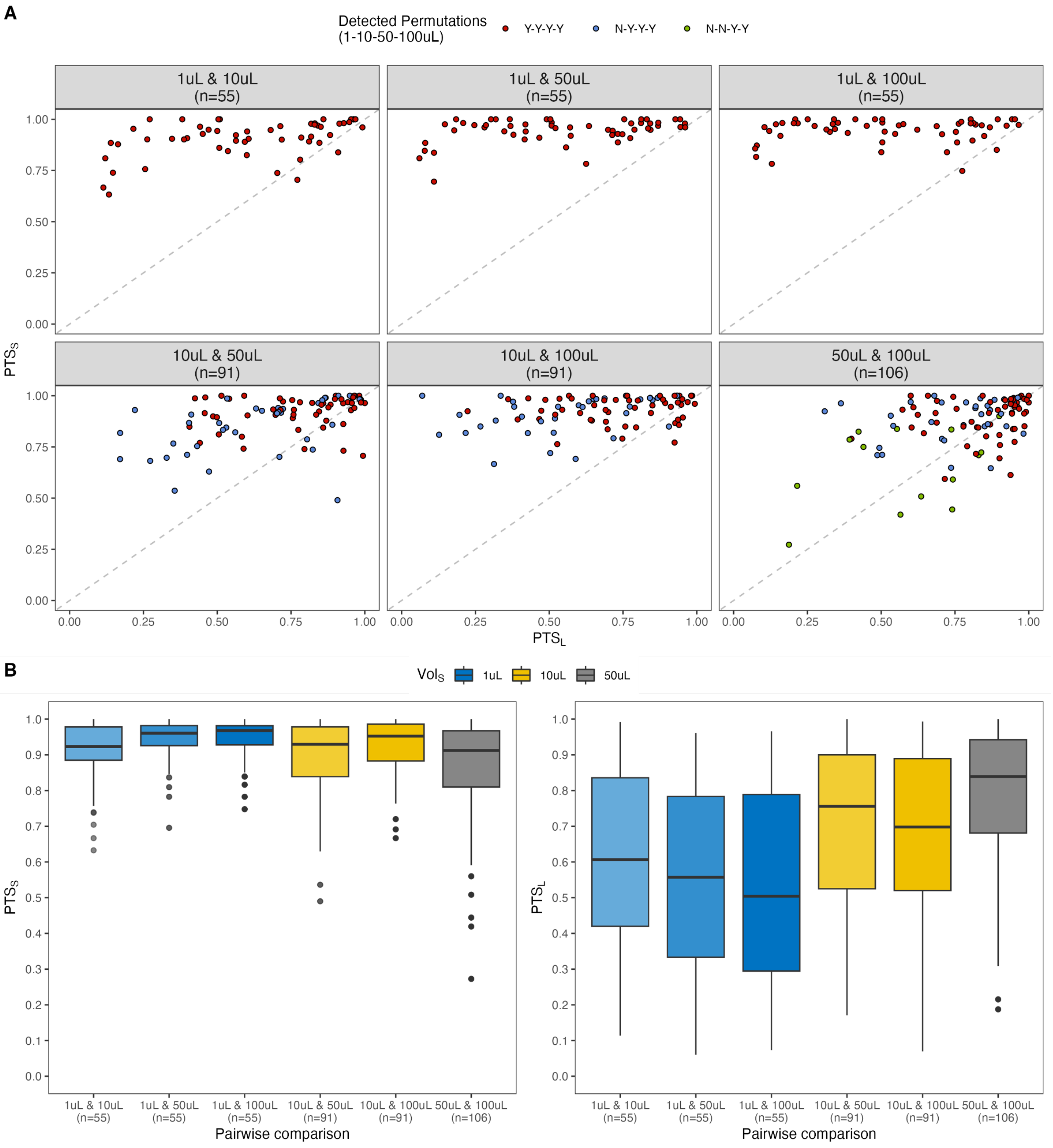

**Fig. S4. Directional genetic similarity between pairwise sampling of pRBC volumes, coloured by (A) *Plasmodium* spp. parasite density, and (B) age group of isolates (i.e. host).** Scatterplots are shown for isolates with Pf-MOI ≥ 1 for all four pRBC volumes (and with isolate repertoire size ≥ 20). $PTS_S$ and $PTS_L$ represent directional genetic similarity levels calculated relative to repertoire sizes of the smaller (S) and larger (L) pRBC volumes, respectively, in a pairwise comparison. Each data point represents an isolate. In (A), data points without density information are coloured in grey.

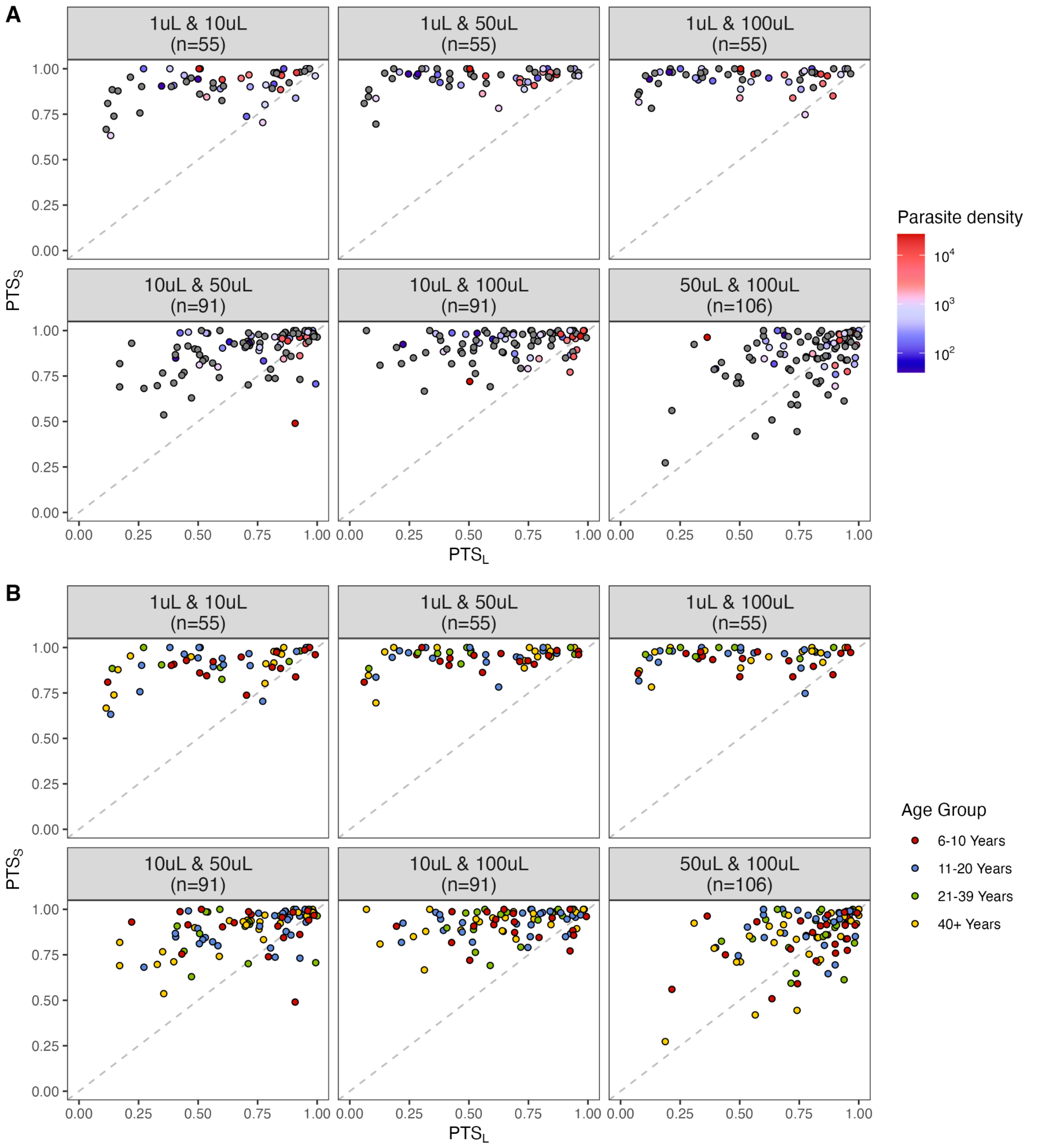

**Fig. S5. Analysis of repeat isolates in the context of varying pRBC volumes.** Here, “R1” represents 40 field isolates included in the initial dataset while “R2” represents the repeat of these isolates. **(A)** Estimated Pf-MOI for the repeat of every isolate, represented on the y-axis. Both the initial (R1) and repeat (R2) isolates confirmed the volume-based observations reported in this study, showing increased detection of *P. falciparum* and Pf-MOI for both R1 and R2 sets of isolates as larger pRBC volumes were sampled. **(B)** Comparison of genetic similarity by PTS between repeats at every pRBC volume. PTS values were lowest for repeats of 1µL pRBC volumes and highest for the largest 100µL pRBC volume, suggesting that sampling consistency was greater when done in larger volumes. **(C)** Based on Lin’s concordance correlation coefficient (CCC), there was strong concordance in estimated Pf-MOI between repeats of all pRBC volumes, particularly in larger pRBC volumes of 50µL or 100µL. **(D)** Distributions of isolate repertoire size and Pf-MOI showed no significant differences between repeats (adjusted p-value > 0.05). Statistical analysis was conducted with the pairwise Wilcoxon signed rank tests. Levels of significance in pairwise tests were adjusted using the “holm” correction. Median Pf-MOI for both R1 and R2 increased from 1 to 2 comparing the 1µL and 100µL pRBC datasets.

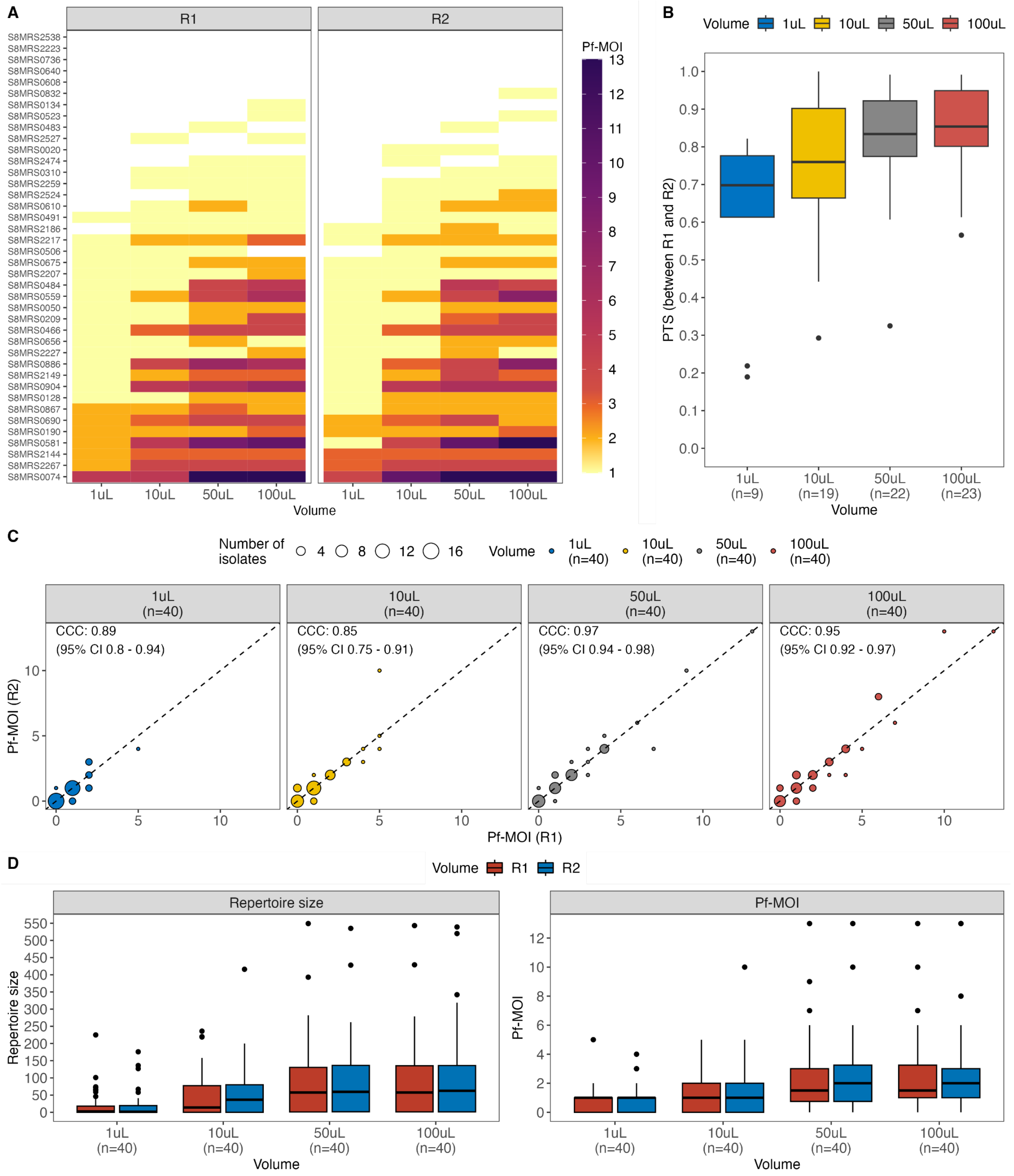

**Fig. S6. Comparison of Pf-MOI values for isolates with Pf-MOI = 0 for one pRBC volume but Pf-MOI > 0 in a different pRBC volume.** Pf-MOI$_S$ and Pf-MOI$_L$ represent the Pf-MOI values of the smaller and larger compared pRBC volumes, respectively. For such isolates, the estimated Pf-MOI for two pRBC volumes mostly differed by one.

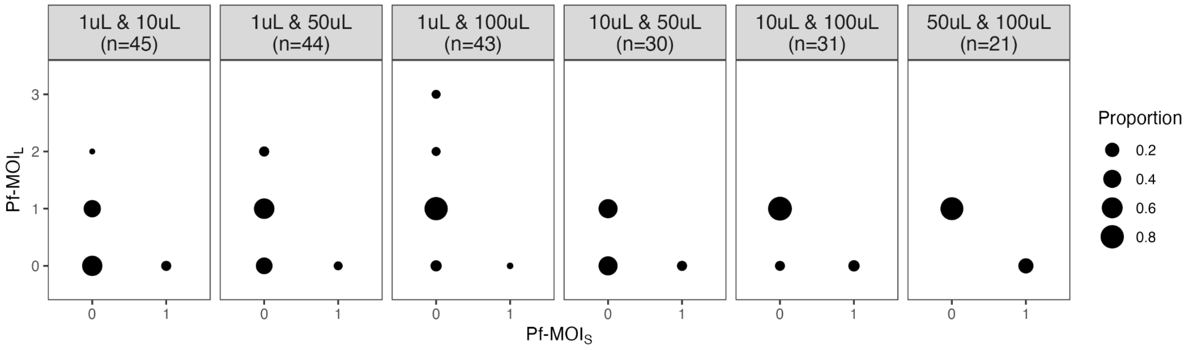

**Fig. S7. Non-linear relationships between fold differences in Pf-MOI (FD$_{Pf\text{-}MOI}$, shown in log scale) and the Pf-MOI in smaller compared pRBC volumes (Pf-MOI$_S$).** Scatterplots are shown for isolates with Pf-MOI ≥ 1 (and with isolate repertoire size ≥ 1) for all four pRBC volumes. FD$_{Pf\text{-}MOI}$ is shown in log scale on the y-axis. Horizontal red, dashed line at value of $2^0$ indicates no difference in estimated Pf-MOI (FD$_{Pf\text{-}MOI}$ = $2^0$ = 1). Green solid lines show fitted smooth curves, grey background indicates the 95% CI.

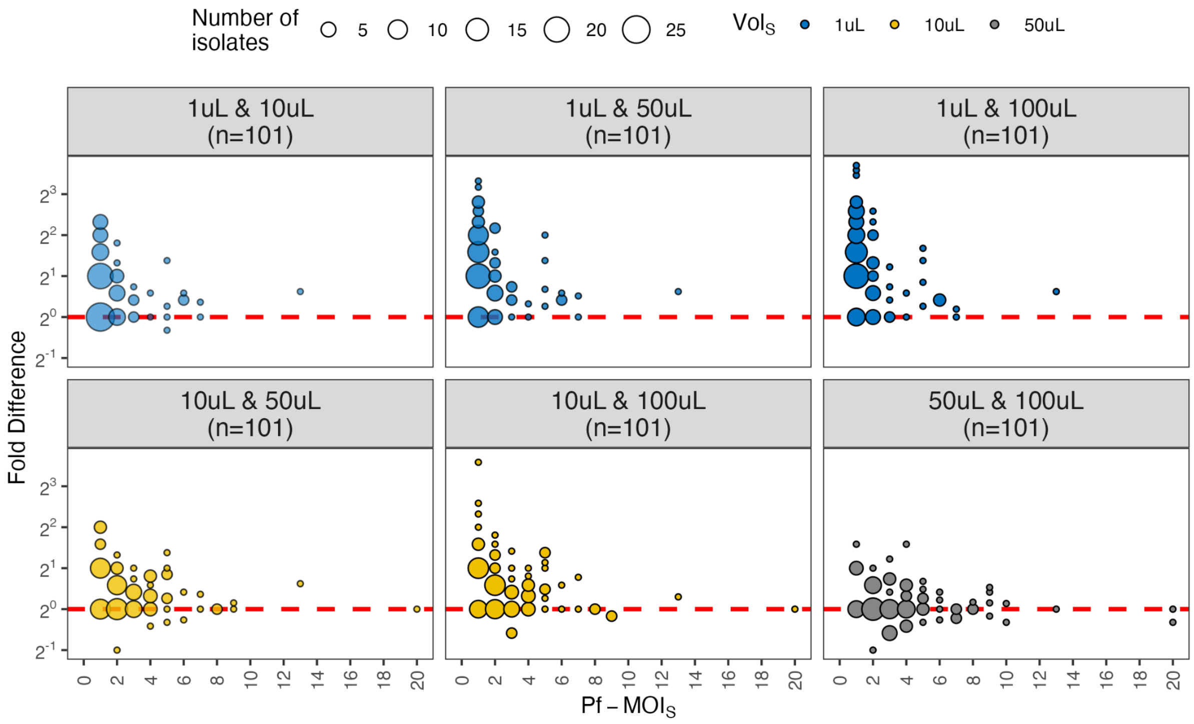

**Table S5. Power calculation based on proportion of samples with Pf-MOI > 1.** This estimates statistical power (p) of the current dataset to provide correction scales between 1µL and larger pRBC volumes (α = 0.05). This was then followed by estimations of sample sizes (n) required to achieve sufficient power (power ≥ 0.80, α = 0.05) for correction scales stratified by host age group.

| CURRENT DATASET | | | | | | | | | Sample size (n) required to achieve p ≥ 0.80 |
|---|---|---|---|---|---|---|---|---|---|
| Host age group (years) | pRBC vol 1 | pRBC vol 2 | n1[+] (vol1) | n2[+] (vol2) | % Pf-MOI>1[^] (vol1) | % Pf-MOI>1[^] (vol2) | Effect size (h) | Statistical power | |
| All | 10µL | 1µL | 101 | 101 | 66.7 | 32.5 | 0.78 | 1.00 | |
| | 50µL | 1µL | 101 | 101 | 81.2 | 32.5 | 1.16 | 1.00 | |
| | 100µL | 1µL | 101 | 101 | 85.5 | 32.5 | 1.29 | 1.00 | |
| 6-10 | 10µL | 1µL | 24 | 24 | 74.2 | 51.6 | 0.56 | 0.50[#] | 50 |
| | 50µL | 1µL | 24 | 24 | 77.4 | 51.6 | 0.68 | 0.65[#] | 34 |
| | 100µL | 1µL | 24 | 24 | 87 | 51.6 | 0.99 | 0.93 | 16 |
| 11-20 | 10µL | 1µL | 29 | 29 | 82.1 | 46.4 | 0.73 | 0.79[#] | 30 |
| | 50µL | 1µL | 29 | 29 | 96.4 | 46.4 | 1.14 | 0.99 | 13 |
| | 100µL | 1µL | 29 | 29 | 92.9 | 46.4 | 1.02 | 0.97 | 16 |
| 21-39 | 10µL | 1µL | 21 | 21 | 62.5 | 16.7 | 1.11 | 0.95 | 13 |
| | 50µL | 1µL | 21 | 21 | 70.8 | 16.7 | 1.34 | 0.99 | 9 |
| | 100µL | 1µL | 21 | 21 | 79.2 | 16.7 | 1.46 | 1.00 | 8 |
| ≥40 | 10µL | 1µL | 27 | 27 | 50 | 14.7 | 0.87 | 0.89 | 21 |
| | 50µL | 1µL | 27 | 27 | 79.4 | 14.7 | 1.57 | 1.00 | 7 |
| | 100µL | 1µL | 27 | 27 | 82.4 | 14.7 | 1.86 | 1.00 | 5 |

[+] Sample sizes available for two compared pRBC volumes.

[^] Proportion of samples with MOI > 1 (%).

[#] Comparisons where power < 0.80 at a significance level (α) of 0.05.

**Table S6. Estimated genetic similarity between isolate repertoires by pairwise type sharing (PTS) categorised by host age group and sampled pRBC volumes.** These estimates were based on isolates with Pf-MOI ≥ 1 (and with isolate repertoire size ≥ 20).

| **Population metrics** | **1µL** | **10µL** | **50µL** | **100µL** |
|---|---|---|---|---|
| Number of isolates | 55 | 91 | 106 | 113 |
| DBLα type richness | 4,682 | 8,486 | 10,760 | 11,715 |
| upsA | 611 | 903 | 1,062 | 1,126 |
| non-upsA | 4,071 | 7,583 | 9,698 | 10,589 |
| Pairwise type sharing | | | | |
| Minimum | 0.000 | 0.000 | 0.000 | 0.000 |
| Maximum | 0.124 | 0.301 | 0.400 | 0.365 |
| Mean | 0.018 | 0.026 | 0.028 | 0.027 |
| Median | 0.017 | 0.024 | 0.026 | 0.026 |
| Q1 | 0.000 | 0.014 | 0.016 | 0.015 |
| Q3 | 0.027 | 0.035 | 0.038 | 0.038 |

**Fig. S8. Rarefaction curves of DBLα tags in the upsA and non-upsA groups.** These estimates were based on isolates with Pf-MOI ≥ 1 (and with isolate repertoire size ≥ 20).

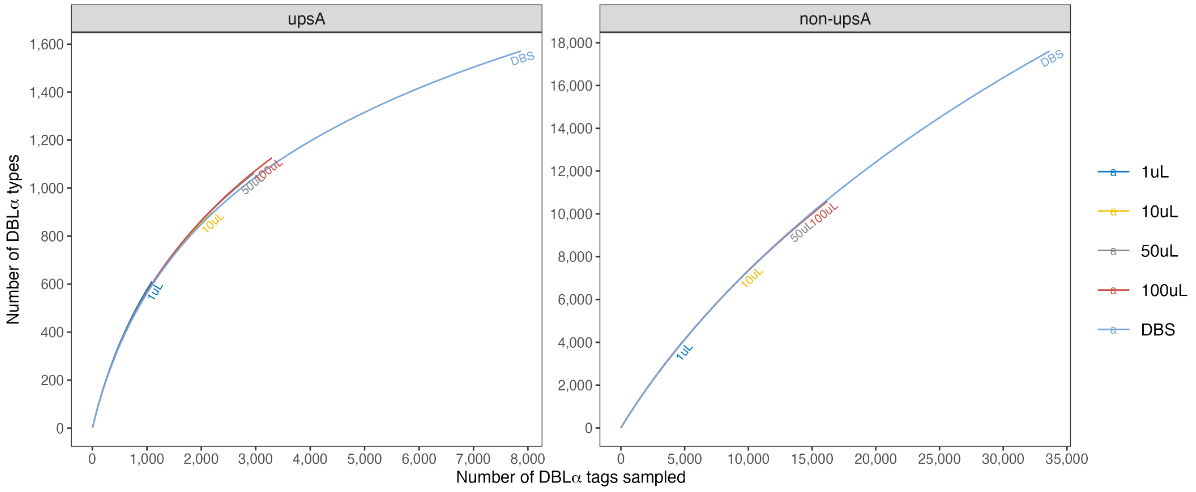

**Fig. S9. Proportion of DBLα types recovered from DBS and/or pRBC samples.** These estimates were based on isolates with Pf-MOI ≥ 1 (and with isolate repertoire size ≥ 20). This analysis included DBS samples (263 isolates) and pRBC samples (55, 91, 106, and 113 isolates for 1µL, 10µL, 50µL, and 100µL volumes, respectively). Each data point represents the proportion of DBLα types in the reservoir identified in isolates with specific combinations of DBS and pRBC volume (values in Table S7). E.g. for 100µL pRBC, of the 21,222 DBLα types identified in 263 DBS and 113 pRBC samples, 32.1% was present in both "**DBS & pRBC**", 23.1% in "**pRBC only**", and the remainder of 44.8% in "**DBS only**". Finding subsets of DBLα types exclusively from different sample sources suggests that both sampling approaches are necessary and complementary to more accurately reflect true population-level metrices.

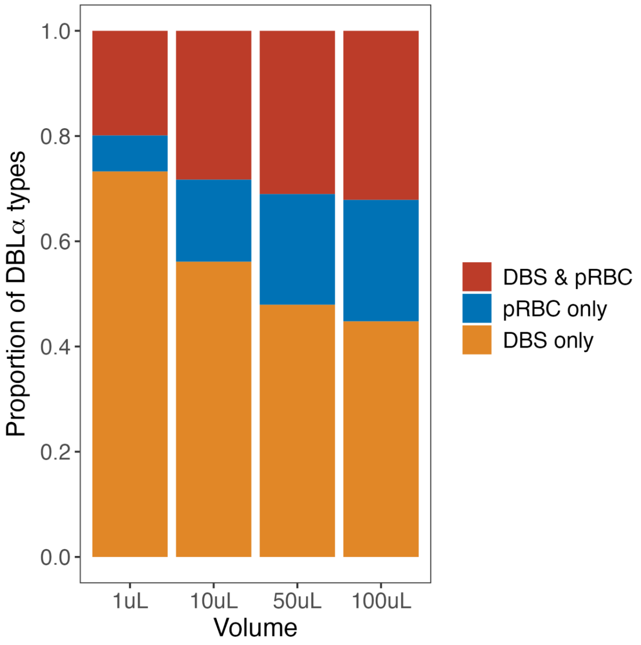

**Table S7. Proportion of DBLα types recovered from DBS and/or pRBC samples.** These estimates were based on isolates with Pf-MOI ≥ 1 (and with isolate repertoire size ≥ 20).

| pRBC volume | Combination | Number of DBLα types | Proportion of DBLα types (%) |
|---|---|---|---|
| 1µL | DBS & pRBC | 3,486 | 19.9% |
| | pRBC only | 1,196 | 6.8% |
| | DBS only | 12,842 | 73.3% |
| 10µL | DBS & pRBC | 5,472 | 28.3% |
| | pRBC only | 3,014 | 15.6% |
| | DBS only | 10,856 | 56.1% |
| 50µL | DBS & pRBC | 6,415 | 31.0% |
| | pRBC only | 4,345 | 21.0% |
| | DBS only | 9,913 | 48.0% |
| 100µL | DBS & pRBC | 6,821 | 32.1% |
| | pRBC only | 4,894 | 23.1% |
| | DBS only | 9,507 | 44.8% |

**Table S8. Statistical tests to examine association between cumulative complexity and host/spatial characteristics.** Human host characteristics include sex (male/female), age (years), haemoglobin levels, axillary temperature (°C), and occupation. Spatial characteristics include village (Vea/Gowrie) and sections.

| Parameters | p-value | Statistic | Estimate | Statistical test |
|---|---|---|---|---|
| ***Host characteristics*** | | | | |
| Sex | 0.436 | W = 2434 | NA | Wilcoxon Rank Sum Test |
| Age | 0.421 | S = 542,268.42 | -0.067 (rho) | Spearman Rank Correlation Test |
| Haemoglobin levels | 0.199 | S = 453,552.77 | 0.107 (rho) | Spearman Rank Correlation Test |
| Axillary temperature | 0.644 | S = 478,337.70 | 0.039 (rho) | Spearman Rank Correlation Test |
| Occupation | 0.843 | $\chi^2$ = 2.72 | NA | Kruskal-Wallis Rank Sum Test |
| ***Spatial characteristics*** | | | | |
| Village | 0.773 | W = 2,535 | NA | Wilcoxon Rank Sum Test |
| Section | 0.950 | $\chi^2$ = 4.57 | NA | Kruskal-Wallis Rank Sum Test |

**Table S9. Adjusted prevalence of different *Plasmodium* spp. was used to predict the number of missed cases in the Bongo District and the Upper East Region in Ghana when using only DBS data.** Fold difference in the observed prevalence from DBS and 100µL in this study was first estimated, i.e. "× difference". Infection prevalence was then estimated from DBS data from a larger surveyed group of *N*=1,809 individuals living in Bongo who were sampled at the same time, i.e. "Survey DBS - Bongo", stratified by age groups of 6-9 years (*n*=230), 10-19 years (*n*=563), 20-39 years (*n*=271), and ≥40 years (*n*=391). This was further adjusted through multiplication with the "× difference" factor to obtain prevalence in Bongo with deeper sampling, i.e. "Adjusted". Infection prevalence was then used to estimate the number of *Plasmodium* spp. infections in the Bongo District and the Upper East Region of Ghana, based on total and age-structured population size data accessed through the City Population website based on the 2021 Population and Housing census by the Ghana Statistical Service (GSS).

| Age Groups (as per City Population) | Prevalence (This Study, *N*=188) | | | Number of infections (Survey DBS, *N*=1,455) | | Prevalence (Survey DBS, *N*=1,455) | | Number of infections (Bongo, *N*=102,004) | | Number of infections (Upper East, *N*=1,100,676) | |
|---|---|---|---|---|---|---|---|---|---|---|---|
| | DBS | 100µL | × difference | DBS | Adjusted | DBS | Adjusted | DBS | Adjusted | DBS | Adjusted |
| ***P. falciparum*** | | | | | | | | | | | |
| All ages (≥6 years) | 55.85 | 73.94 | 1.32 | | | | | | | | |
| 6-9 years | 40.63 | 62.50 | 1.54 | 107 | 165 | 46.52 | 71.57 | 5,660 | 8,707 | 62,200 | 95,692 |
| 10-19 years | 62.07 | 81.03 | 1.31 | 381 | 497 | 67.67 | 88.35 | 19,283 | 25,175 | 205,798 | 268,680 |
| 20-39 years | 46.81 | 65.96 | 1.41 | 147 | 207 | 54.24 | 76.43 | 19,058 | 26,855 | 205,560 | 289,653 |
| ≥40 years | 66.67 | 80.39 | 1.21 | 213 | 257 | 54.48 | 65.69 | 14,278 | 17,217 | 154,664 | 186,506 |
| ***P. malariae*** | | | | | | | | | | | |
| All ages (≥6 years) | 7.45 | 14.36 | 1.93 | | | | | | | | |
| 6-9 years | 6.25 | 9.38 | 1.50 | 11 | 16 | 4.78 | 7.17 | 582 | 873 | 6,394 | 9,592 |
| 10-19 years | 15.52 | 27.59 | 1.78 | 106 | 188 | 18.83 | 33.47 | 5,365 | 9,537 | 57,256 | 101,788 |
| 20-39 years | 2.13 | 8.51 | 4.00 | 5 | 20 | 1.85 | 7.38 | 648 | 2,593 | 6,992 | 27,967 |
| ≥40 years | 3.92 | 7.84 | 2.00 | 11 | 22 | 2.81 | 5.63 | 737 | 1,475 | 7,987 | 15,975 |
| ***P. ovale* spp.** | | | | | | | | | | | |
| All ages (≥6 years) | 2.13 | 5.32 | 2.50 | | | | | | | | |
| 6-9 years | 3.13 | 9.38 | 3.00 | 5 | 15 | 2.17 | 6.52 | 264 | 793 | 2,907 | 8,720 |
| 10-19 years | 3.45 | 6.90 | 2.00 | 31 | 62 | 5.51 | 11.01 | 1,569 | 3,138 | 16,745 | 33,489 |
| 20-39 years | 0.00 | 2.13 | 3.13 | 3 | 9 | 1.11 | 3.46 | 389 | 1,216 | 4,195 | 13,121 |
| ≥40 years | 1.96 | 3.92 | 2.00 | 8 | 16 | 2.05 | 4.09 | 536 | 1,072 | 5,809 | 11,618 |
| ***P. vivax*** | | | | | | | | | | | |
| All ages (≥6 years) | 0.00 | 0.00 | 0.00 | 0 | 0 | 0.00 | 0.00 | 0 | 0 | 0 | 0 |
