## Supplementary Data 2 for "Metagenomic analysis reveals extreme complexity of *Plasmodium* spp. infections in high transmission in West Africa"

### Estimation of whole blood (WB) and packed red blood cell (pRBC) equivalent in dried blood spots (DBS)

Blood samples were obtained with consent from a male in their 30's (Hb 11.8 g/dL, RDT negative). First, we pipetted specific volumes of blood in triplicates (1, 2, 5, 10, 15, 20, 25, 30, 50µL) onto 3MM Whatman filter papers. We then let these DBS air dry as we would in the field for ~45min to 1 hour. We then measured the diameter of each DBS across two directions (*d1* and *d2*) and calculated the average diameter (*d*) for each DBS across the two measurements. The average radius (*r*) values were used to calculate the area (*A*) of DBS (Area, $A = \pi r^2$) (Supplementary Data 2.1).

**Supplementary Data 2.1. Measurements of DBS diameters and calculations of average DBS area**

| WB Volume (µL) | Repeat | DBS Diameter (d1, cm) | DBS Diameter (d2, cm) | Average DBS Diameter (d, cm) | Average DBS Radius (r, cm) | Average DBS Area (A, cm$^2$) |
|---|---|---|---|---|---|---|
| **1.00** | 1 | 0.40 | 0.30 | 0.35 | 0.18 | 0.10 |
| | 2 | 0.30 | 0.30 | 0.30 | 0.15 | 0.07 |
| | 3 | 0.40 | 0.40 | 0.40 | 0.20 | 0.13 |
| **2.00** | 1 | 0.50 | 0.50 | 0.50 | 0.25 | 0.20 |
| | 2 | 0.40 | 0.50 | 0.45 | 0.23 | 0.16 |
| | 3 | 0.50 | 0.50 | 0.50 | 0.25 | 0.20 |
| **5.00** | 1 | 0.70 | 0.70 | 0.70 | 0.35 | 0.38 |
| | 2 | 0.80 | 0.70 | 0.75 | 0.38 | 0.44 |
| | 3 | 0.70 | 0.60 | 0.65 | 0.33 | 0.33 |
| **10.00** | 1 | 0.80 | 0.90 | 0.85 | 0.43 | 0.57 |
| | 2 | 0.80 | 0.80 | 0.80 | 0.40 | 0.50 |
| | 3 | 0.90 | 0.90 | 0.90 | 0.45 | 0.64 |
| **15.00** | 1 | 1.10 | 1.00 | 1.05 | 0.53 | 0.87 |
| | 2 | 1.00 | 1.00 | 1.00 | 0.50 | 0.79 |
| | 3 | 1.00 | 1.10 | 1.05 | 0.53 | 0.87 |
| **20.00** | 1 | 1.20 | 1.10 | 1.15 | 0.58 | 1.04 |
| | 2 | 1.10 | 1.20 | 1.15 | 0.58 | 1.04 |
| | 3 | 1.10 | 1.10 | 1.10 | 0.55 | 0.95 |
| **25.00** | 1 | 1.30 | 1.30 | 1.30 | 0.65 | 1.33 |
| | 2 | 1.20 | 1.20 | 1.20 | 0.60 | 1.13 |
| | 3 | 1.30 | 1.20 | 1.25 | 0.63 | 1.23 |
| **30.00** | 1 | 1.40 | 1.30 | 1.35 | 0.68 | 1.43 |
| | 2 | 1.30 | 1.20 | 1.25 | 0.63 | 1.23 |
| | 3 | 1.30 | 1.20 | 1.25 | 0.63 | 1.23 |
| **50.00** | 1 | 1.50 | 1.50 | 1.50 | 0.75 | 1.77 |
| | 2 | 1.50 | 1.40 | 1.45 | 0.73 | 1.65 |
| | 3 | 1.40 | 1.50 | 1.45 | 0.73 | 1.65 |

This resulted in a non-linear relationship between WB volume (µL) and the DBS area (cm$^2$), shown below (Supplementary Data 2.2).

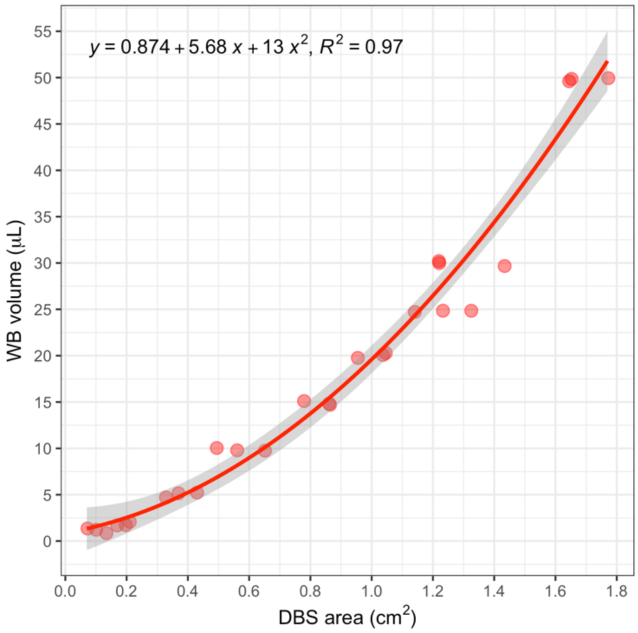


**Supplementary Data 2.2. Relationship between WB volume on DBS and area of DBS.**

Based on this relationship, we estimated the volume of blood we obtain when we cut (based on area):

i. One 0.5cm × 0.5cm square (A = 1 × 0.25cm$^2$) → ~3.11µL of WB
ii. Two 0.5cm × 0.5cm squares (A = 2 × 0.25cm$^2$) → ~6.21µL of WB
iii. One 0.5cm × 1 cm rectangle (A = 1 × 0.50cm$^2$) → ~6.97µL of WB

DBS cuttings in our previous studies have used methods ii and iii[1,2], giving us an average of ~6-7µL equivalent of WB volume. Given the average expected proportion of RBCs at about 40%[3], this then translates into the equivalent of ~2.4-2.8µL packed red blood cells (pRBC).

**Additional notes:** We noted that, the DBS were larger when using larger volumes of blood, as was expected. However, more importantly, larger blood volumes tended to soak further through into the filter paper. In contrast, smaller volumes of blood expanded but did not soak through the filter as much compared to the larger blood volumes. This explains the non-linear relationship between WB volume collected on the DBS and the calculated DBS area.
