## Supplementary Data 1 for "Metagenomic analysis reveals extreme complexity of *Plasmodium* spp. infections in high transmission in West Africa"

Detailed material and methods for this study

### 1. Study area and population

This study was conducted in the Bongo District (Vea/Gowrie catchment area) located in the Upper East Region of northern Ghana, characterised by seasonal malaria transmission, with a short, wet season (Jun-Oct) and a prolonged dry season (Nov-May)[1,2]. Before this research was undertaken, informed consent was sought and obtained from the key stakeholders and the local community in Bongo District. In addition, members of the local community were trained as field workers and were directly involved in liaising with the local community and in the collection of the study data. At the time of this survey, the population had undergone malaria control interventions, including long-lasting insecticidal nets (LLINs), three rounds of indoor residual spraying (IRS) from 2013 to 2015, as well as five consecutive years (2016-2020) of seasonal malaria chemoprevention (SMC) administered at monthly intervals during the malaria season to children between the ages of 3-59 months (i.e. < 5years old)[3]. This study was reviewed and approved by the ethics committees at the Navrongo Health Research Centre, Ghana (NHRC IRB-131) and The University of Melbourne, Australia (HREC 21649).

### 2. Estimation of WB and pRBC volume equivalent in DBS

The typical protocol in the field involves collecting three to four small, dried blood spots (DBS) per filter paper[4]. From these small DBS, two sections are cut (approximately 5mm × 5mm each) for gDNA extraction. We estimated the average volume of whole blood in these DBS by pipetting specific volumes of blood from a male in their 30's (Hb 11.8 g/dL, RDT negative) onto 3MM Whatman filter papers in triplicates, then measuring the average DBS diameter (cm) and calculating the average DBS area ($cm^2$). The resulting non-linear relationship between DBS volume (µL) and DBS area ($cm^2$) approximated that two 5mm × 5mm DBS cuttings would contain the equivalent of ~6 to 7µL of whole blood (Supplementary Data 2). Given the average expected proportion of RBCs at about 40%[5], our typical DBS cuttings would thus have the equivalent of ~2.4 to 2.8µL packed red blood cells (pRBC), with some variation due to a person's hermatocrit levels. Other notable sources of variability in this estimation include the finger prick process and the pressing of blood to the filter paper, as well as hemoglobin (Hb) levels will also affect the DBS size and how blood soaks into the filter paper. Thus, this study sets out to compare MOIs estimated using gDNA extracted from pRBC volumes of 1µL, 5µL, 10µL, and 100µL.

### 3. Parasitological measurements

Parasitological measurement methods were performed as previously published[6,7]. Briefly, parasite densities were counted against 200 white blood cells (WBC) on 10% Giemsa-stained thick film blood smears and examined under oil immersion of 100-fold magnification. Final parasite densities (parasites per mL of blood) were calculated by averaging two independent readings, assuming an average WBC count of 8,000 per µL of blood. Parasite species were identified using a 100-fold magnification of thin film smears and categorised based on morphology.

### 4. Sampling of venous WB and DBS samples

#### *4.1. Whole blood (WB)*

A study population of 200 individuals with ages ranging from 6 to 90 years were sampled. Children <5 years currently receiving seasonal malaria chemoprevention (SMC) were excluded. Approximately 5mL of venous whole blood samples (WB) were collected per person in EDTA tubes, temporally stored on ice packs (2°C to 8°C), and then transported to the laboratory at the Navrongo Health Research Centre, Ghana, where they were processed within 2 to 4 hours after collection in the field. These WB samples were centrifuged at 3,000 RPM at 4°C for 10 minutes to separate the WB into components of packed red blood cells (pRBC), white blood cells, and plasma. All three components were harvested individually and stored frozen at -80°C.

#### *4.2. Dried blood spots (DBS)*

At the same time as the venous whole blood collection, DBS were also collected onto filter paper (3MM Whatman) for the same participants and for a larger subset of the population (see [3] for detailed methods on DBS collection).

### 5. Extraction of genomic DNA from pRBC and DBS

#### *5.1. From packed red blood cells (pRBC)*

Of the 200 individuals (i.e. “isolates”), a total of 12 were excluded (Table S1). Nine were excluded as these could not be matched accurately to epidemiological data. The remaining three isolates were excluded as these were later confirmed to be symptomatic (i.e. febrile and microscopically-positive for *P. falciparum*). Genomic DNA was extracted from four volumes of pRBC (1µL, 10µL, 50µL, and 100µL) from the remaining 188 afebrile isolates, using the QIAamp DNA Blood Mini Kit, following manufacturer's protocol (Cat No. 51106). 1× PBS was added to pRBC samples to make up 200µL of every sample volume for extraction. An RNase A step was included to generate RNA-free genomic DNA, following manufacturer's protocol (Cat No. 19101). Genomic DNA was eluted in 50µL buffer AE and stored at -80°C. In subsequent sections, a "sample" refers to a DBS or pRBC volume per isolate, hence there can be multiple samples per isolate.

#### *5.2. From dried blood spots (DBS)*

Two 5mm × 5 mm sections were cut from DBS and gDNA extraction was performed using the QIAamp DNA Mini Kit (Cat No. 51306) according to manufacturer's protocol with modifications[1]. Genomic DNA was eluted in 50µL buffer AE and stored at -80°C.

### 6. Measurement of PfHRP2 concentrations in plasma

Plasma PfHRP2 concentration was determined using the Quantimal CELISA kit (TM, Cellabs). In this enzyme-linked immunosorbent assay, wells are coated with a primary monoclonal antibody to PfHRP2, sample is applied and wells are washed, then probed with a secondary anti-Pf antibody conjugated to horseradish peroxidase (HRP) for detection using the FLUOstar omega plate reader (BMG Labtech) at 450nm. A standard curve was prepared starting at 10ng/mL of standardised PfHRP2 and diluted 1:2 down to 0.01ng/mL. The negative control of Melbourne plasma (Australian Red Cross Lifeblood, non-malarious region) was kindly provided by Prof Stephen Rogerson's lab for comparison. Participant plasma samples were collected as mentioned above, and diluted 1:20 in duplicate, along with the negative control and RPMI blanks, to allow most positive samples to fall within the range of detection of the assay. Where the sample was saturated it was repeated at 1:40, and where it was detected but fell under the cut-off it was repeated at 1:10 to confirm results. All standard curves and samples were analysed using Arigo GAINdata ELISA data analysis software (Arigo biolaboratories) to obtain PfHRP2 concentrations, and the cut-off was applied manually at the level suggested by the manufacturer (OD negative control+0.1, Cellabs).

### 7. Detection of *Plasmodium* spp. using *18S rRNA* species-specific PCR

*Plasmodium* species detection was performed on the DBS and 100µL pRBC samples (*N*=188 isolates). To detect the presence of different *Plasmodium* species, previously published protocols with modifications were used[1,8]. A nested polymerase chain reaction (nPCR) targeting the 18S ribosomal RNA gene (*18S rRNA*) was performed to identify *Plasmodium* species in DBS and pRBC samples, as previously done[1]. All PCR reactions, both the first and second rounds, were carried out in a total volume of 20µL consisting of 4µL of 5x buffer, 1.6µL of 25mM $MgCl_2$, 0.25µL of 10mM dNTPs, 1µL of 2.5µM of each primer, and 0.08µL of GoTaq G2 Flexi DNA polymerase (Promega, Cat No. M7805). Using 2µL of extracted gDNA as template, the first round of PCR included amplification of the *Plasmodium 18S rRNA* with genus-specific forward and reverse primers (rPLU5 and rPLU6). Subsequently, using 2µL aliquots of PCR product from the first PCR, the second PCR was performed with species-specific primer pairs (rFAL1/rFAL2) to detect presence of *P. falciparum*, (rMAL1/rMAL2) for *P. malariae*, (rOVA1/rOVA2) for *P. ovale* spp., (rVIV1/rVIV2) for *P. vivax*[8]. The following cycling conditions were used, as previously described method[1]: Round 1: 95ºC for 2 min, 25 cycles of 58ºC for 2 min, 72ºC for 5 min and 94ºC for 1 min, followed by 58ºC for 2 min ad 72ºC for 2 min; and Round 2: 95°C for 2 min, 30 cycles of 58°C for 2min, 72°C for 5min and 94°C for 1min, followed by 58°C for 2min and 72°C for 2min. As quality control, positive (*P. falciparum* 3D7 isolate) and negative controls were included. Visualization was made on a 2% agarose gel stained with SYBR™ Safe DNA Gel Stain (Invitrogen) along with a 100 bp Ladder DNA marker (100 bp to 3,000 bp) (Axygen). The different *Plasmodium* spp. were determined to be present if a band at 205 bp was observed for *P. falciparum*, 144 bp for *P. malariae*, 800 bp for *P. ovale* spp., and 120 bp for *P. vivax*.

### 8. Genotyping of *P. malariae* microsatellites

Of the 188 isolates, 26 were identified as *P. malariae*-infected isolates through species-specific PCR. The 100µL pRBC sample for each of these isolates was genotyped at 12 neutral microsatellite markers using primers previously published[9,10]. The semi-nested PCR protocol used is adapted for low-density infections (Rios-Teran et al., 'In preparation'). The first round of PCR involves amplifying with four sets of primers in three reactions. Each first-round reaction was carried out in a total volume of 25 µL, comprising 5 µL of genomic DNA, 5 µL of

5× buffer, 3 µL of 25 mM $MgCl_2$, 1 µL of 10 mM dNTPs mix, 0.4 µL of 2.5 µM of each primer, and 0.24 µL of GoTaq G2 Hot Start polymerase (Promega), and free nuclease water. The second round required the preparation of twelve reactions with the fluorescent-labelled primers. Each reaction contained same quantities of buffer and $MgCl_2$ than the first round, and 3µL of the PCR product from the first reaction with 0.5µL of 10 mM dNTPs mix, 0.8µL of 2.5µM of each primer, 0.12µL of GoTaq G2 Hot Start polymerase (Promega), and free nuclease water up to 25µL. The cycling conditions for the first round were as follows: 4 minutes at 94°C, 35 cycles of 30 seconds at 94°C, 30 seconds at 48°C, and 1 minute at 68°C, the final extension was at 68°C for 2 minutes and storage at 4°C. For the second round the cycles were increased to 45 with 30 seconds at 52°C of annealing temperature. For the genotyping procedure, four pools were prepared per isolate to be analysed for fragment analysis by capillary electrophoresis at AGRF. Allele sizes were measured by comparing with the size standard LIZ500. The data was then uploaded to GeneMarker® software v3.0.1 from SoftGenetics to score the alleles. Only the alleles that were greater than one third of the dominant allele were scored to avoid stutter peaks. Automated binning was performed using *tandem*[11].

### 9. Identification and genotyping of *P. ovale* spp. using *potra*

Of the 188 isolates, 10 were identified as *P. ovale* spp.-infected isolates through the *18S rRNA* species-specific PCR. The 100µL pRBC sample for each of these isolates was genotyped for *P. ovale* spp. identification (i.e., *P. ovale curtisi and P. ovale wallikeri*) and subsequently genetic diversity. This semi-nested PCR protocol involves *P. ovale* spp. specific amplification of a size-polymorphic fragment of the tryptophan-rich antigen gene (*potra*)[12,13] with modifications for low-density infections (Rios-Teran et al., 'In preparation'). For the first reaction, the primers (PoTRA-F/PoTRA rev3) target regions conserved between the two species. The total volume was 20µL for the first reaction, and it contained 4µL of 5x buffer, 1.6µL of 25mM $MgCl_2$, 0.25µL of 10mM of dNTP mix, 1µL of 2.5µM of each primer, 0.08µL of GoTaqG2 Flexi DNA polymerase, and 2µL of DNA as template. For the secondary reaction, the forward PoTRA-F primer was used in two separate reactions per isolate, but with reverse primers specific to each of the two *P. ovale* spp., either PocTRA-R for *P. ovale curtisi* or PowTRA-R for *P. ovale wallikeri*. The secondary reaction used the same reagent quantities but with 3µL of the product of the first reaction. The cycling conditions for the first round of the semi-nested PCR started with 4 minutes at 95°C, followed by 25 cycles of 1 minute at 95°C, 1 minute at 56°C, and 1 minute at 72°C, the final extension was at 72°C for 2 minutes and storage at 4°C. For the second round the cycles were 30 with 1 minute at 60°C of annealing temperature. Storage is at 4°C as well. The product of the secondary reaction (10µL) was electrophoresed for two hours on a 2% agarose gel stained with SYBR™ Safe DNA Gel Stain (Invitrogen) along with a 100 bp Ladder DNA marker (100 bp to 3,000 bp) (Axygen). The gels were visualized under Image Lab software BIORAD Gel Doc™ EZ Imager version 6.1. The species of *P. ovale curtisi* or *P. ovale wallikeri* was determined to be present if a band was seen around 400 bp to 600 bp. The molecular weights were compared with the DNA ladder, recorded in Microsoft Excel v. 2211, and later analysed in RStudio version 4.2.2.

### 10. Targeted amplicon sequencing of *P. falciparum* DBLα tags with *var*coding

The *var*coding method has been shown to be appropriate for MOI estimation in high transmission, outperforming SNP-based barcodes[14,15]. This method utilises targeted amplification and sequencing of the *var* DBLα type sequences encoding the Duffy-binding-like domain of *Plasmodium falciparum* erythrocyte membrane protein 1 (PfEMP1). For DBS samples, *var*coding was performed on microscopy-positive isolates using primers and protocols described in [3]. A modified *var*coding protocol was applied to the four pRBC volumes:

#### *10.1. Isolates, Repeats, and Controls*

Four pRBC volumes for 188 isolates were *var*coded (i.e. genotyped at the DBLα tag region), including those that appear negative by microscopy or species-specific PCR, in order to assess if more infections are found when larger volumes are sampled. Forty isolates (i.e. ~20% of isolates) were randomly selected as repeats to assess the reproducibility of *var*coding outcomes. Laboratory strains (i.e. 3D7, Dd2, HB3) were included as positive controls. Samples of varying pRBC volumes of a same isolate and its repeats (if selected as repeat) were sequenced in the same sequencing pool and run.

#### *10.2. PCR primers*

As described[3], the sequence region within *var* genes encoding the DBLα domain of PfEMP1 (i.e. DBLα tags) were amplified in a single-step PCR from genomic DNA using universal degenerate primer sequences to blocks D (forward primer: DBLαAF, 5'-GCACGMAGTTTYGC-3') and H (reverse primer: DBLαBR, 5'-GCCCATTCSTCGAACCA-3')[16,17]. In this study, primers used included forward barcoded primers and an unbarcoded reverse primer. Both forward and reverse primers also contain Illumina Nextera overhangs to facilitate subsequent preparation of sequencing libraries. Primer sequences are available online on GitHub (see Data Availability section).

#### *10.3. PCR*

Each PCR reaction was prepared in a total volume of 40µL with final concentrations of: 0.5X buffer, 2mM of $MgCl_2$, 0.07mM of dNTPs, 0.375µM of each primer (DBLαAF, DBLαBR), 3 units of GoTaq G2 Flexi DNA polymerase (Promega, Cat No. M7805), and 2µL of genomic DNA template. The cycling conditions were as follows: initial denaturation step of 2 min at 95°C was followed by 30 cycles of: 40 seconds at 95°C, 90 seconds at 49°C, 90 seconds at 65°C, and a final extension step of 10 min at 65°C.

#### *10.4. Purification, Quantification, Pooling, and Sequencing*

The PCR products were purified using the SPRI method (solid-phase reversible immobilization) (Agencourt AMPure XP beads (Cat No. A63881)). Purified PCR product concentrations were measured using the Quant-iT PicoGreen dsDNA Kit (Invitrogen, Cat No. P7589) following manufacturer's protocol. Amplicons were then pooled equimolarly, with each pool consisting of up to 99 isolates, all with unique MID tags. Pooled amplicons were subsequently indexed (Amplicon Indexing Service) and sequenced on the Illumina MiSeq platform (2 × 300bp) at the Australian Genome Research Facility (AGRF).

### 11. Processing DBLα tags into DBLα types

An established bioinformatics workflow[18,19] consisting of a suite of pipelines was used to generate DBLα tags from raw Illumina paired-end reads (https://github.com/UniMelb-Day-Lab/tutorialDBLalpha). DBLα tag sequence data included DBLα tags from pRBC generated in this study, combined with DBLα tags from an interrupted time-series study in Bongo, involving one pilot and eight cross-sectional surveys conducted between 2012 to 2020 (i.e. Malaria Reservoir Study). These surveys involved mostly asymptomatic isolates[1,3,20] with small proportions of symptomatic and clinical isolates (unpublished). Clustering of DBLα tags was performed at a 96% nucleotide identity threshold [21] used to define a DBLα tag in a sample or DBLα types in a population of tags.

- The DBLaCleaner pipeline (v1.0)[18] was used to generate DBLα tag sequences per sample from raw paired-end Illumina sequence reads.
- The clusterDBLa pipeline (v1.0)[18] was used to generate unique DBLα types from DBLα tags with a matrix detailing presence/absence of each DBLα type in isolates.
- The classifyDBLa pipeline (v1.0)[19] was used to classify DBLα types into DBLα domain subclasses and subsequently into upsA or non-upsA groups as per [22].

### 12. Estimation of isolate repertoire size and multiplicity of infection (Pf-MOI)

The isolate repertoire size represents the number of unique DBLα types in an isolate. Multiplicity of infection (Pf-MOI) represents the estimated number of unique parasite genomes in an isolate. For samples with ≥20 DBLα types, multiplicity of infection in each sample was estimated using a Bayesian approach (prior="uniform", aggregate="pool")[3]. Samples with ≥1 DBLα types but <20 DBLα types were assigned Pf-MOI=1. Pm-MOI was estimated based on the maximum number of distinct alleles at any of the 12 microsatellite loci. Po-MOI was estimated from the number of unique fragment sizes obtained based on the *potra* gene.

### 13. Estimation of fold difference (FD) and model fitting

Differences in isolate repertoire sizes and Pf-MOI were expressed as fold differences (FD):

$$FD_{size} = \frac{Size_L}{Size_S}; FD_{MOI} = \frac{MOI_L}{MOI_S}$$

where $Size_S$ and $Size_L$ are the isolate repertoire sizes of smaller (*S*) and larger (*L*) volume samplings, respectively, of a same isolate, and $\text{Pf-MOI}_S$ and $\text{Pf-MOI}_L$ are the Pf-MOI of the smaller (*S*) and larger (*L*) volume samplings, respectively, of a same isolate. A generalised additive model (GAM) was fit to the data ($FD_{Pf\text{-}MOI}$ and $\text{Pf-MOI}_S$) with penalised smoothing parameters selected by REML (*gam* function in mgcv v1.9-1[23]).

### 14. Estimation of genetic similarity or overlap between isolate repertoires

To ensure robust analyses exploring genetic similarity, we evaluated only samples with ≥ 20 DBLα types (i.e. isolate repertoire size ≥ 20). Genetic similarity of repertoires of two isolate repertoires is generally represented by the pairwise type sharing metric (PTS)[21]. Specifically:

$$PTS = \frac{2 * shared_{ij}}{Size_i + Size_j}$$

where $shared_{ij}$ is the shared number of DBLα types between isolate repertoires of samples *i* and *j*, and $Size_i$ and $Size_j$ are the isolate repertoire sizes of samples *i* and *j*, respectively. A value of 0 indicates the absence of sharing between two samples while a value of 1 indicates completely identical var DBLα repertoires.

Genetic similarity between isolate repertoires from two different pRBC volumes was assessed in a directional manner relative to the isolate repertoire size of a reference sampled volume (i.e. denominator) [18]:

$$PTS_S = \frac{shared_{SL}}{Size_S}; PTS_L = \frac{shared_{SL}}{Size_L}$$

where $shared_{SL}$ is the shared number of DBLα types between isolate repertoires of smaller (*S*) and larger (*L*) volume samplings of a same isolate, and $Size_S$ and $Size_L$ are the isolate repertoire sizes of smaller and larger volume samplings, respectively, of a same isolate (i.e. $Vol_S$ and $Vol_L$). High $PTS_S$ and low $PTS_L$ values indicate that DBLα types identified in the smaller volume are also present in the larger volume and that there are additional DBLα types identified in the larger volume but not found in the smaller volume.

### 15. Adjusting prevalence, Pf-MOI, and parasite census population size with scale up to population sizes in Ghana

#### 15.1. Prevalence

We estimated prevalence of the different species based on DBS from 1,809 individuals living in Bongo collected at the time of this study. Prevalence was calculated by species (*P. falciparum*, *P. malariae*, *P. ovale* spp.) and age (6-9 years, 10-19 years, 20-39 years, ≥40 years). Prevalence levels were further adjusted based on the fold differences observed between DBS or 100µL in Fig 1, estimated in this study with *18S rRNA* with the same age stratification.

#### 15.2. Mean MOI and parasite census population size

We first estimated Pf-MOI for 247 individuals living in Bongo. These individuals were microscopy-positive for *P. falciparum* with Pf-MOI ≥ 1 based on *var*coding of DBS samples.

Using the GAM model fitted to the 1µL *vs* 100µL data (see section 1.13 above), these DBS-estimated Pf-MOI values were further adjusted to predict Pf-MOI values that would be measured from deeper sampled volume. Mean MOI was calculated for both DBS and adjusted values without age stratification. Subsequently, this was used to calculate parasite census population size[24]:

$$parasite\ census\ pop\ size = mean\ MOI * prevalence * host\ population\ size$$

### 15.3. Scaling up to larger population sizes

Prevalence, mean MOI, and parasite census population size values were further scaled up to the larger population sizes of the Bongo District and the Upper East Region of Ghana to obtain an approximation of the number of infections or parasites underestimated in these regions.

Total and age-structured population size data was first accessed through the City Population website[25] based on the 2021 Population and Housing census by the Ghana Statistical Service (GSS)[26]. Population sizes in Ghana:

- Bongo District: 120,254 [total]; 30,416 [0-9 years]; 28,494 [10-19 years]; 35,135 [20-39 years]; 26,209 [≥40 years].
- Upper East Region: 1,301,226 [total]; 334,250 [0-9 years]; 304,105 [10-19 years]; 378,958 [20-39 years]; 283,913 [≥40 years].

Given the exclusion of children ≤ 5 years from this study, we further split the population size data for 0-9 years with a 6:4 ratio into two age groups 0-5 years and 6-9 years. Relevant to our study. The final age-structured population size data as follows:

- Bongo District: 102,004 [≥6 years]; 12,166 [6-9 years]; 28,494 [10-19 years]; 35,135 [20-39 years]; 26,209 [≥40 years].
- Upper East Region: 1,100,676 [≥6 years]; 133,700 [6-9 years]; 304,105 [10-19 years]; 378,958 [20-39 years]; 283,913 [≥40 years].

Map of regions and districts in Ghana in Fig 4A was drawn with Global ADMinistrative area (GADM) data downloaded for Ghana (v4.1)[27].

### 16. Statistical analysis

Differences in the number of *Plasmodium* spp. species detected between DBS and 100µL pRBC were analysed with Fisher's exact test (*fisher.test* in *stats* v4.2.1[28]). The nonparametric Friedman rank sum test (*friedman_test* in *rstatix* v0.7.2[29]) was used to compare isolate repertoire sizes and MOI distributions for paired data of four pRBC volumes. Further, pairwise Wilcoxon signed rank tests with "holm" correction (*pairwise_wilcox_test* in *rstatix* v0.7.2[29]) was used to compare paired data of two pRBC volumes or paired data of two repeats for a same isolate. The Lin's concordance correlation coefficient (CCC) values were calculated (*CCC* function in *DescTools* v0.99.49[30]) to estimate the agreement of isolate repertoire sizes and MOI values between two different pRBC volumes for a same isolate or between two repeats of a same isolate. In the repeat analysis, the Kruskal-Wallis Rank Sum test (*kruskal_test* in *rstatix* v0.7.2[29]) was performed to compare PTS distributions between pairs of repeats for each pRBC volumes, followed by Dunn's test (*dunn_test* in *rstatix* v0.7.2[29]) for multiple comparisons. Tests with *p-value* or adjusted *p-value* ≤ 0.05 were considered significant.

To obtain age-specific correction scales, we must have sufficient data with MOI > 1 in each pRBC volume and/or host age groups. To determine the minimum sample sizes to achieve enough statistical power, we first calculated effect sizes (h) from the proportion of isolates with MOI > 1 using the *ES.h* function in the *pwr* R package (v1.3-0)[31]. The *pwr.2p2n.test* function from the same R package was used to estimate statistical power at a significance level (α) of 0.05 (alternative="two.sided"). The *pwr.2p.test* function was further used to estimate the minimum sample sizes (n) required to achieve a statistical power of 0.80 (α=0.05, alternative="two.sided") for future correction by host age groups.

Association of variables with cumulative complexity was tested using the Mann-Whitney U test (for qualitative variables with 2 groups, i.e. sex, village), the Kruskal-Wallis rank sum test (for qualitative variables with >2 groups, i.e. occupation, section), and Spearman's rank correlation (for quantitative variables, i.e. age, haemoglobin, axillary temperature). Tests with *p-value* ≤ 0.05 were considered significant.
